# Typological distinction of remotely sensed metrics of neighborhood vegetation for environmental health intervention design

**DOI:** 10.1101/2023.03.03.23286763

**Authors:** Daniel Fleischer, Jay Turner, Ivan Heitmann, Brent Bucknum, Aruni Bhatnagar, Ray Yeager

## Abstract

The extent to which urban vegetation improves environmental quality and affects the health of nearby residents is dependent on typological attributes of “greenness”, such as canopy area to alleviate urban heat, grass to facilitate exercise and social interaction, leaf area to disperse and capture air pollution, and biomass to absorb noise pollution. The spatial proximity of these typologies to individuals further modifies the extent to which they impart benefits and influence health. However, most evaluations of associations between greenness and health utilize a single metric of greenness and few measures of proximity, which may disproportionately represent the effect of a subset of mediators on health outcomes.

To develop an approach to address this potentially substantial limitation of future studies evaluating associations between greenness and health, we measured and evaluated distinct attributes, correlations, and spatial dependency of 13 different metrics of greenness in a residential study area of Louisville, Kentucky, representative of many urban residential areas across the Eastern United States. We calculated NDVI, other satellite spectral indices, LIDAR derived leaf area index and canopy volume, streetview imagery derived semantic view indices, distance to parks, and graph-theory based ecosystem connectivity metrics. We utilized correlation analysis and principal component analysis across spatial scales to identify distinct groupings and typologies of greenness metrics.

Our analysis of correlation matrices and principal component analysis identified distinct groupings of metrics representing both physical correlates of greenness (trees, grass, their combinations and derivatives) and also perspectives on those features (streetview, aerial, and connectivity / distance). Our assessment of typological greenness categories contributes perspective important to understanding strengths and limitations of metrics evaluated by past work correlating greenness to health. Given our finding of inconsistent correlations between many metrics and scales, it is likely that many past investigations are missing important context and may underrepresent the extent to which greenness may influence health. Future epidemiological investigations may benefit from these findings to inform selection of appropriate greenness metrics and spatial scales that best represent the cumulative influence of the hypothesized effects of mediators and moderators. However, future work is needed to evaluate the effect of each of these metrics on health outcomes and mediators therein to better inform the understanding of metrics and differential influences on environments and health.

## Introduction

Multiple observational studies have found that neighborhood vegetation is associated with improved health outcomes (James et al., 2015,Barboza et al., 2021). However, most studies that examine links between greenness and health rely on greenness metrics such as canopy and NDVI, with little consideration or exploration of other metrics of greenness. Given the many moderators through which greenness affects health, specific greenness metrics may be disproportionately associated with some moderators and subsequent health outcomes.

Various metrics of neighborhood vegetation exposure have been employed in health studies, including Normalized Difference Vegetation Index (NDVI), land-cover classifications (such as tree canopy area), residence distance to parks or other greenspace, leaf area index, street-level green-view index and surveys of perceived greenness, although the most common metric has been NDVI. NDVI is calculated from remote sensing platforms measuring reflected near-infrared and visible portions of the electromagnetic spectrum. Plant photosynthesis consumes visible wavelength photons (with peaks in the red and blue spectral regions), while re-radiating near-infrared photons. Thus, plant photosynthetic area appears bright in the near-infrared and dark in the red portions of the spectrum of a multispectral sensor. NDVI is the ratio normalized to be between -1 and 1:

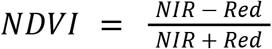

Generally, Tall evergreen trees and deciduous trees at the peak of foliation typically have NDVI values from 0.5 to 0.9, while shrubs and grasses can be in the range of 0.2 to 0.5 (Gamon et al., 1995). Bare soil or rock is generally below 0.1 (Bhandari et al., 2012), while surface water is typically around 0.0 (Buma, 2012). These categorizations are highly fluid; correlations between vegetation characteristics and NDVI can vary significantly depending on sensor configurations, atmospheric effects and phenology, and can exhibit rapid changes due to water and nutrient status. In urban environments, well maintained lawns of manicured turf grass may have NDVI nearly indistinguishable from nearby tall trees. (Caturegli et al., 2016) conducted a detailed assessment of drone acquired (high spatial-resolution) NDVI from turfgrass and found that it varied between 0.5 and 0.9, with a consistent positive relationship between NDVI and nitrogen content/application. (Xiong, 2005) found NDVI for bermuda grass to be between 0.65 to 0.9. Consequently, urban planners faced only with positive correlations between NDVI and community health outcomes will be unguided as to the relative merits of planting trees vs fertilizing grass.

It is clear that there is much physical variation within equivalent values of NDVI and canopy, such as plant health, height, surface area, and density. Such variation is likely to have disproportionate effects on the moderators of relationships between greenness on health, including physical activity, social cohesion, stress, urban heat, microbiome diversity, and air pollution. From an intervention design perspective, the structure of proposed interventions is dependent on the proposed mechanism for health improvement. For example, a greening intervention geared towards reducing air pollution might focus on dense, tall roadside vegetation for interception of traffic sourced particulates, a heat island mitigation strategy might focus on canopy-forming trees over pavement, while an intervention to increase physical activity and social cohesion might focus on increasing open grassy areas where people can gather and exercise. At this level of design consideration, the metrics used to establish the link between greenness and health outcomes becomes a driving factor for evidence-based design.

There have been many poignant critiques of NDVI and canopy as a meaningful measure for environmental health (Rugel et al., 2017, Villeneuve et al., 2018, Reid et al., 2018, Trethewey & Reynolds, 2021), with explicit calls for more nuanced and comprehensive greening metrics (Taylor & Hochuli, 2017, Saleh et al., 2019, Rojas-Rueda et al., 2021, Donovan et al., 2022) to inform our understanding of how greenness influences health and subsequent intervention design and implementation. Thus, the purpose of the present study is to determine groupings and redundancy of greenness metrics and identify potential alternative greenness metrics in order to inform and contextualize greenness metrics utilized in health studies for furthering evidence-based intervention design.

## Materials and Methods

### Study area

The area of study encompasses 12 square kilometers of the southern portion of Louisville, Kentucky shown within the purple outline in figure 1. We used this area for all analyses the employ averaging or summing greening metrics within a buffer around residential addresses. An extended study area of 65 square kilometers was used for direct raster to raster analysis (without residential buffers), which is shown in figure ST-1. For all variables the principal study period is the summer of 2019. We chose to focus on this area as it is the subject of the ongoing Green Heart Project. The Green Heart project is a study employing urban vegetation as an intervention in the style of a clinical trial. Starting in the Autumn of 2019, over 3 years a total of 8000 mature evergreen trees were planted throughout a 50 acre target neighborhood. 800 study participants in the target neighborhood and in the surrounding control neighborhood (where no trees were planted) had blood, urine and hair collected and assayed for various biomarkers of cardiovascular health before and 3 years after completing the intervention. Also included in the Green Heart study design is a comprehensive air monitoring program to assess the impact of the new vegetation on various air pollutant concentrations, noise levels, and temperature.

**Figure 1.**
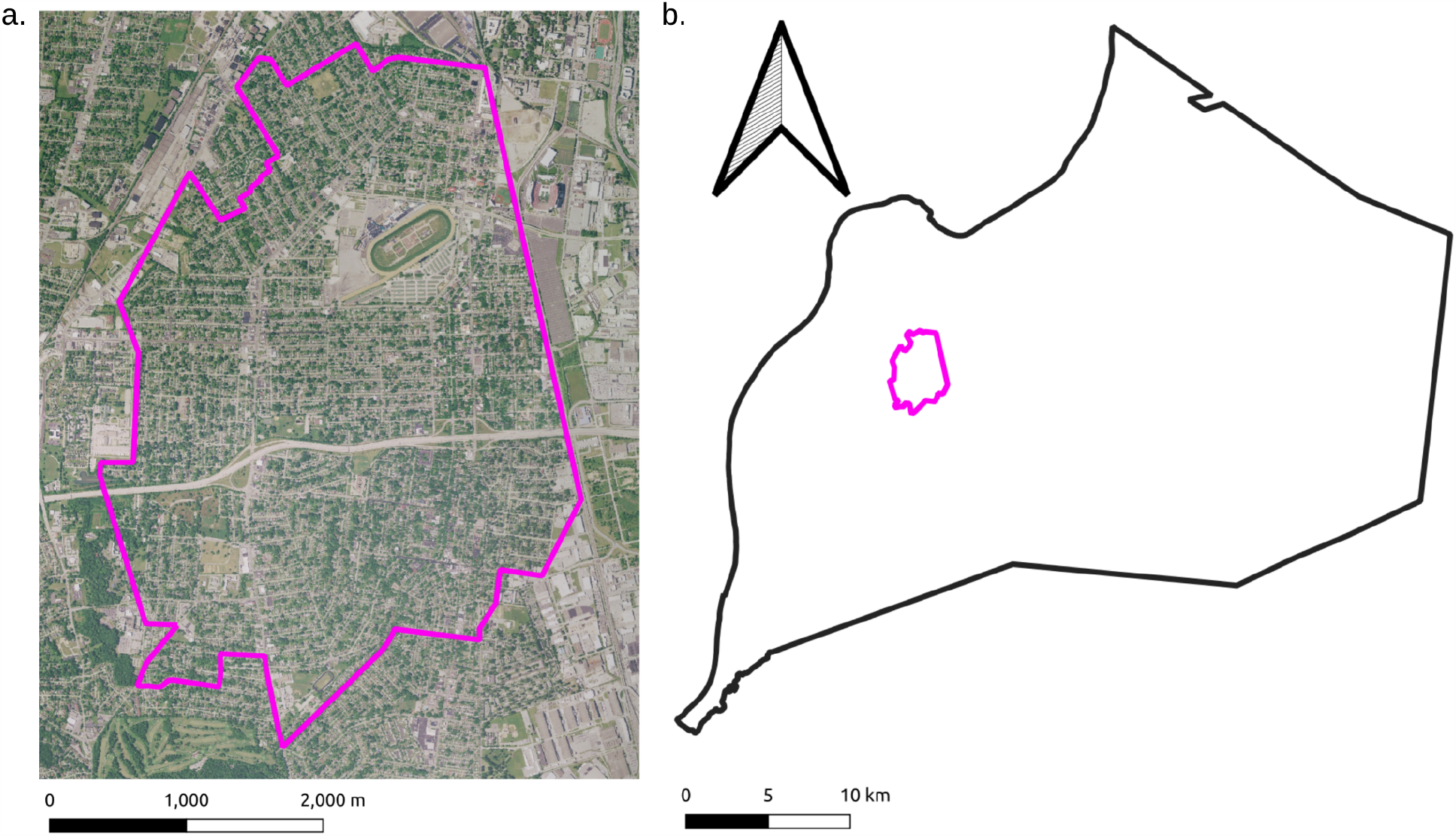
a. Map of study area. b. Study area in context of Jefferson County, KY.

The Green Heart study will provide an opportunity to assess the deterministic relationships between urban vegetation and human health outcomes. However, in order to establish a dose-response, the definition of dosage units must be established, and it may turn out that the vegetation intervention may appear to have more or less powerful effects depending on what methods are used to measure it. By way of a simple example, if the mechanism for health improvement were limited to air pollution mitigation via deposition of particulates on leaf surfaces, then we would expect the association between leaf area index and the health outcomes to be greater than the association between NDVI and the health outcome. Similarly, if the main source of air pollution were traffic sources near roadways, then streetview derived tree metrics might show a stronger association with the health outcome than overall tree cover. More likely there will be multiple mechanisms with intersecting relationships with different greening metrics. For example, increased microbial biodiversity might be associated with health improvement, and also with habitat connectivity, leaf surface area, and plant diversity, and possibly NDVI via its covariance with these other metrics.

### Data utilized

The details of the grenness metrics, their significance, and how they were acquired/calculated is in the supplemental information. A table showing the metrics, their source and very brief description is shown in table 1.

**Table 1:** Greenness metrics used in the present study.

| Metric | Source | Description |
| --- | --- | --- |
| Summer average NDVI | Calculated from Planetscope multispectral 3.5 meter rasters | Normalized Difference Vegetation Index: Measure of healthy vegetation. Can be higher for dense trees, but can also respond similarly to healthy grass. |
| Summer average SAVI | Calculated from Planetscope multispectral 3.5 meter rasters | Soil Adjusted Vegetation Index: Similar to NDVI but adjusted for brightness of bare soil in low vegetation areas. |
| Summer average TDVI | Calculated from Planetscope multispectral 3.5 meter rasters | Transformed Difference Vegetation Index: Similar to SAVI but does not saturate in high vegetation areas. |
| Summer average GCI | Calculated from Planetscope multispectral 3.5 meter rasters | Green Chlorophyll Index: Proxy for chlorophyll content of area. |
| Annual NDVI AUC | Calculated from 18 NDVI measurements throughout 2019 using composite Simpson's rule | NDVI area under the curve: Results from taking NDVI during all seasons to get a phenological curve, and then integrating to get the area under the curve (AUC). |
| LAI | Calculated from aerial LIDAR point cloud using Beer's law | Leaf Area Index: Single sided leaf surface area per unit Earth area. |
| Canopy Volume | Calculated from aerial LIDAR point cloud using top and bottom of canopy height | Total volume of space occupied by tree canopy. |
| Canopy Height Max | Calculated from aerial LIDAR point cloud using top of canopy height | Maximum height of tree canopy |
| Canopy Height Min | Calculated from aerial LIDAR point cloud using bottom of canopy height | Minimum height of tree canopy |
| Canopy Area | Calculated from LAI | Total area under tree canopy |
| Grass area | Calculated from land-cover classification of O'Neil-Dunne & Safavi (2021) | Total area occupied by grass (not under tree canopy). |
| Interaction flux | Calculated from canopy area patches | The local contribution of each habitat patch to the probability of connectivity |
| Corridor-120 | Calculated from canopy area patches | The probable habitat corridors between habitat patches |
| Minimum Distance to Parks | Calculated from address locations and park locations | For each address, the minimum distance between the address and the nearest park. |
| Streetview tree index | Calculated from semantic segmentation of Google street-view images | For each point spaced 20 meters along roadways, the percentage of the spherical viewshed occupied by trees. |
| Streetview grass index | Calculated from semantic segmentation of Google street-view images | For each point spaced 20 meters along roadways, the percentage of the spherical viewshed occupied by grass. |
| Streetview plant index | Calculated from semantic segmentation of Google street-view images | For each point spaced 20 meters along roadways, the percentage of the spherical viewshed occupied by non-tree, non-grass plants. |

In brief the greenness variables that we used in this analysis include the spectral indices, LIDAR derived metrics, streetview derived metrics, and connectivity metrics. The summer averaged spectral indices (summer average NDVI, TDVI, GCI and SAVI) measure the overall area of healthy vegetation (trees, grass and shrubs) during the summer, based largely on the presentation of chlorophyll-based photosystems to the observing satellite. The LIDAR derived metrics include attributes of the tree canopy (Leaf area index (LAI), canopy volume, canopy height maximum, canopy height minimum, canopy area) and grass area. The LAI represents the surface area of tree canopy leaves, which relates to the canopy’s capacity for air pollution capture, evapotranspiration, and biodiversity support. These ecosystem services are also likely to correlate with the canopy volume, area and height metrics. The streetview derived view indices (tree-view, grass-view, plant-view, and total vegetation view) represent the amount of each category of the landscape features visible from roadways, and may relate to the intersection of landscape features with traffic pollution and also common human traversal routes. The connectivity metrics (minimum distance to parks, interaction flux, and habitat corridors) relate to the network properties of the greenness, with interaction flux and habitat corridors relating to the provision of connectivity between habitat patches for microbiota and fauna, and minimum distance to parks relating to accessibility of greenspace to area residents.

The corresponding data *types* broadly fall into 3 categories: continuous raster data, categorical raster data, and point data. The continuous raster data includes all of the spectral indices, the LAI, canopy volume, canopy heights, interaction flux and habitat corridors. The categorical raster data includes the canopy area and grass area, and is at 1 meter spatial resolution, with a value of zero for no canopy or grass, and a value of 1 for canopy or grass. The point data includes all of the streetview data (in points along roadways spaced 20 meters apart) and the minimum distance to parks (calculated for each building address).

The metrics can be ad hoc characterized in a multitude of ways, for example by the physical analyte that they suppose to measure (tree metrics, grass metrics, chlorophyll metrics, graph connectivity metrics), or by the method of their acquisition (spectral indices, LIDAR derived metrics, streetview derived metrics), or they can be categorized objectively using measures of their orthogonality. In this study we use principal component analysis to delineate common dimensions of covariance between the variables, with an eye towards understanding the extent to which each variable is unique or potentially redundant.

### Statistical Analysis

For correlations between raster data and point data, we used the following method: We downloaded all address points within the study area as a geojson file from the LOJIC (Louisville and Jefferson County, KY Information Consortium) open geospatial data website (https://data.lojic.org/). Using Qgis 3.24.3-Tisler we created buffers of 50, 250, 500 and 1000 meters around every address point (shown in figure SI-14). We then used the Python Geopandas library version 0.10.2 to calculate the mean value of each streetview semantic viewshed index falling within each address buffer. We then used the Python Rasterio library version 1.2.10 to calculate the mean value of each continuous raster dataset within each address buffer, and the sum of each categorical raster dataset within each buffer. We then used the Python Pandas library version 1.4.1 to calculate the Spearman correlation matrix for all variables within each set of buffers. We performed principal component analysis and principal component regression using the Python Sci-kit learn library.

For raster to raster comparisons, we used the Python Pandas library to make pixel-to-pixel Spearman’s correlation matrices. Prior to the correlation analysis all rasters were resampled to 4 meter resolution and aligned with the Planetscope datasets using the Qgis raster alignment tool. Pixel-to-Pixel Linear regression residual maps were calculated using GRASS r.regression.multi function in Qgis.

## RESULTS

The Spearman correlation matrix of the variables aggregated into 50 meter buffers around building addresses is shown in figure 2. With the 50 meter buffer, very high correlation (>0.95) is seen between canopy volume, canopy area, and LAI. This is not surprising, as these metrics are all derived from the same LIDAR derived tree canopy dataset. Interaction flux is also derived from this dataset, and shows relatively high correlations of 0.79 to 0.82 with these metrics. These four LIDAR-derived canopy metrics show no correlation with grass area, which is also derived from the LIDAR data, but represents portions of the non-canopy area. Summer NDVI shows a relatively high correlation of around 0.86 with the four LIDAR derived tree canopy metrics, and a similar correlation with annual integrated NDVI AUC, however annual NDVI AUC has a lower correlation of 0.66 with the LIDAR derived tree canopy metrics. This is not surprising in that the south Louisville neighborhood in question has very few evergreen trees, and so the annual NDVI AUC shows a higher score for grassy areas than tree canopy.

**Figure 2.**
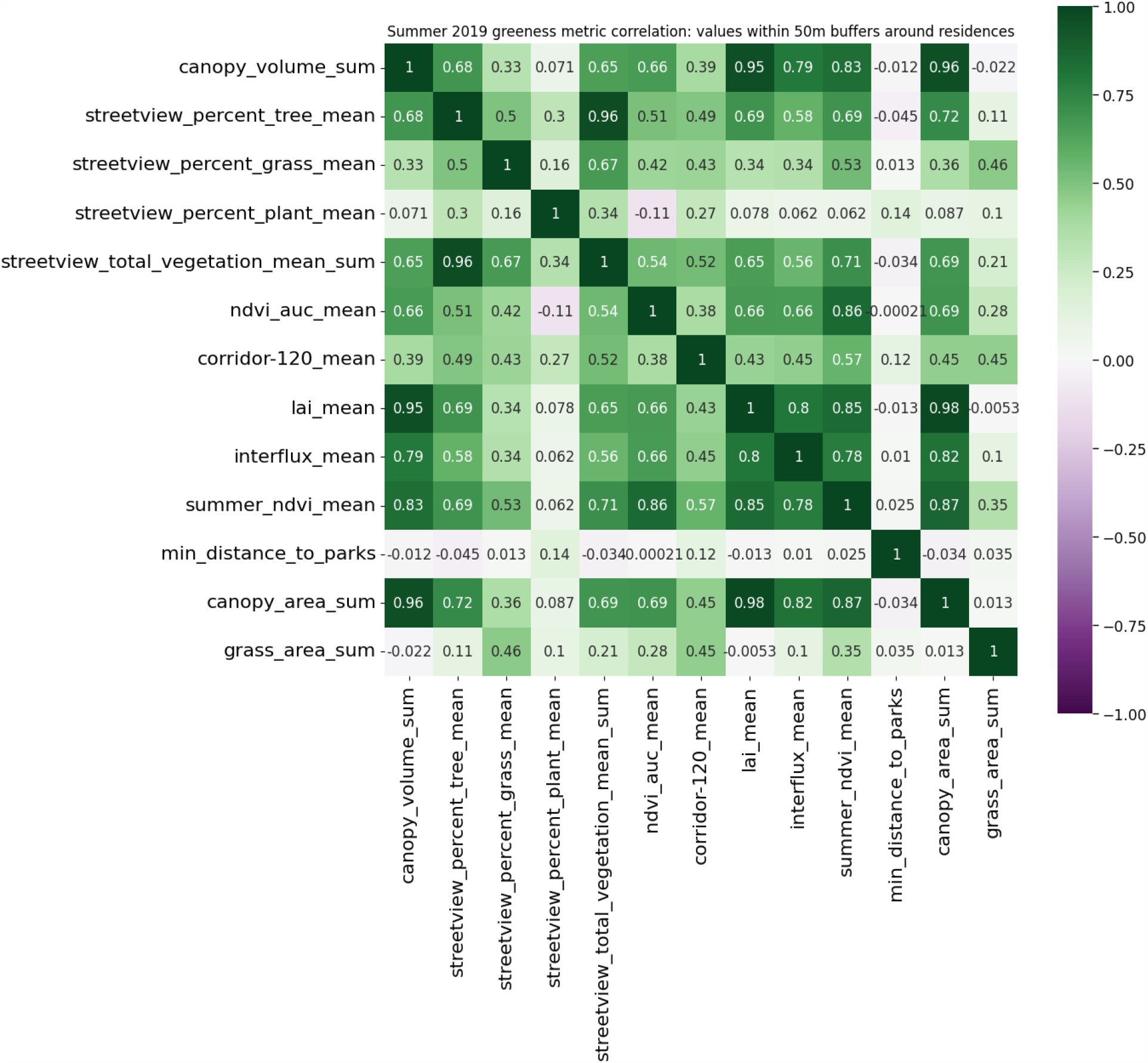
Correlation between mean metrics within a 50 meter buffer around all address points.

The streetview tree index shows correlations between 0.58 and 0.72 with the four LIDAR derived tree canopy metrics, and 0.69 with summer average NDVI. Streetview grass index shows correlations of 0.46, 0.53 and 0.5 with grass area, summer NDVI, and streetview tree index respectively, and lower correlations with the LIDAR derived tree canopy metrics. Streetview plant index and minimum distance to parks both have low or no correlation with most other variables.

Figures 3, 4 and 5 show the correlation matrices using 250, 500 and 1000 meter buffers around address points. What these demonstrate is that as the buffer size increases, all of the metrics see increases in covariance, with the exception of minimum distance to parks.

**Figure 3.**
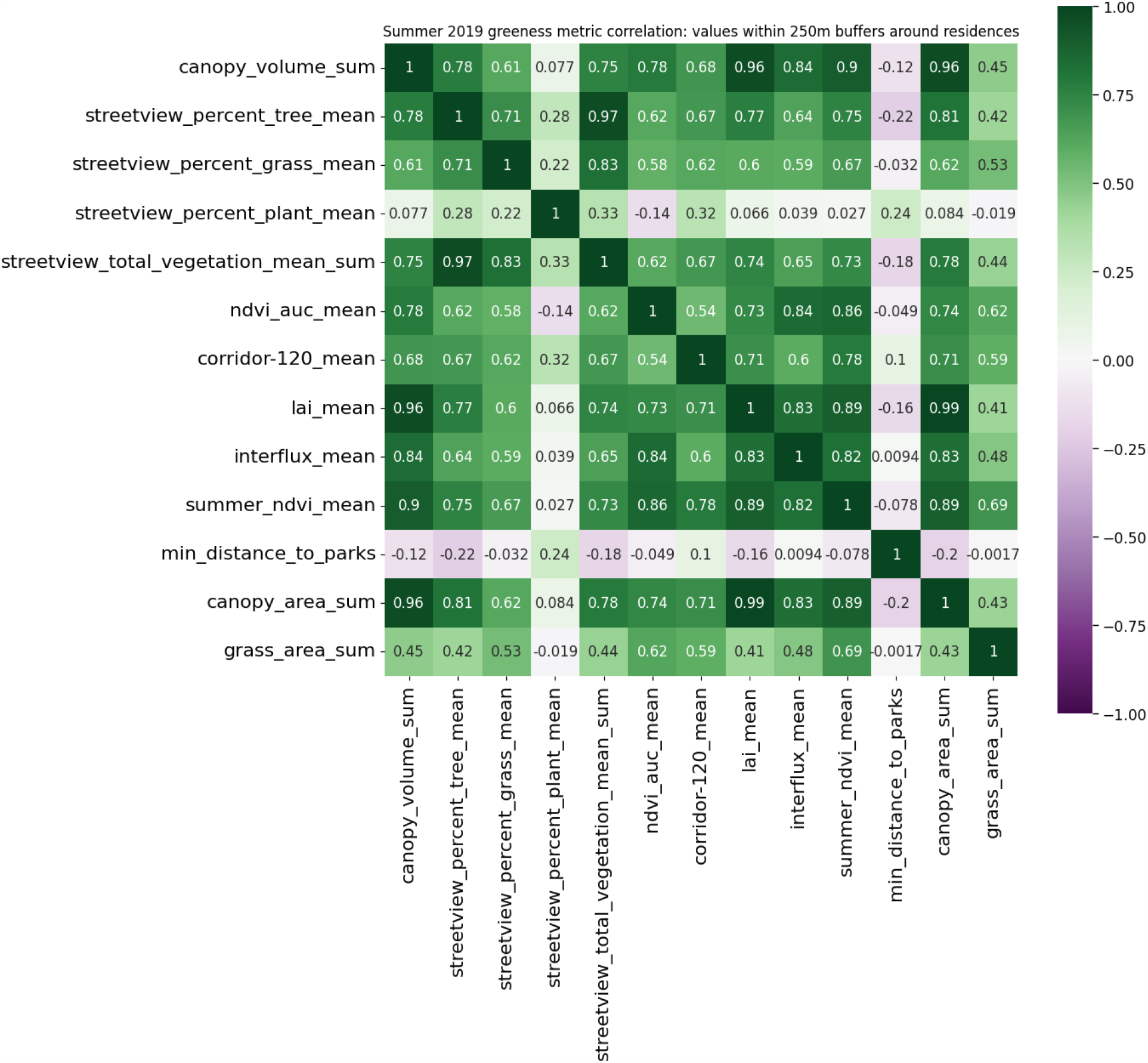
Correlation between mean metrics within a 250 meter buffer around all address points.

**Figure 4.**
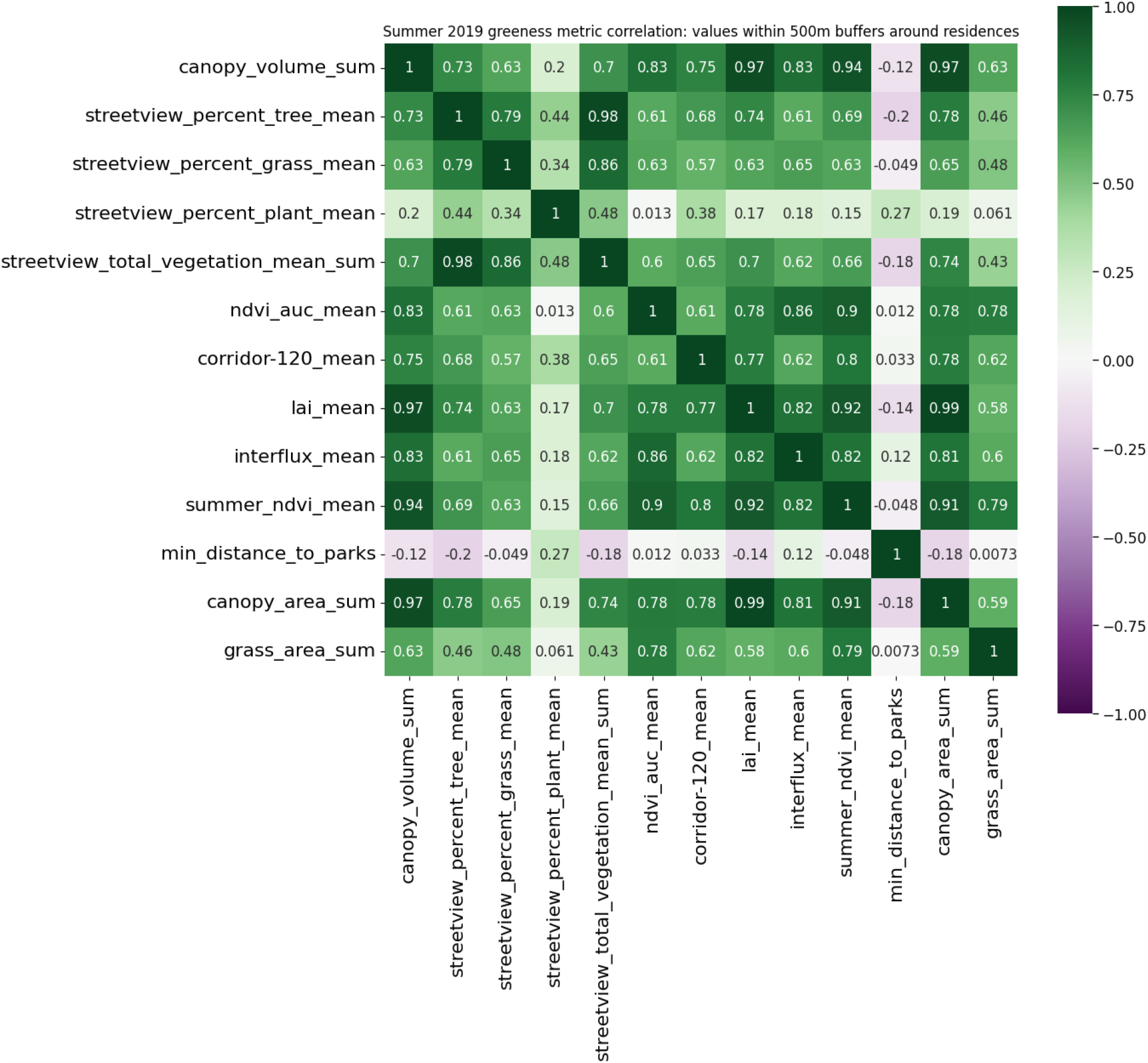
Correlation between mean metrics within a 500 meter buffer around all address points.

**Figure 5.**
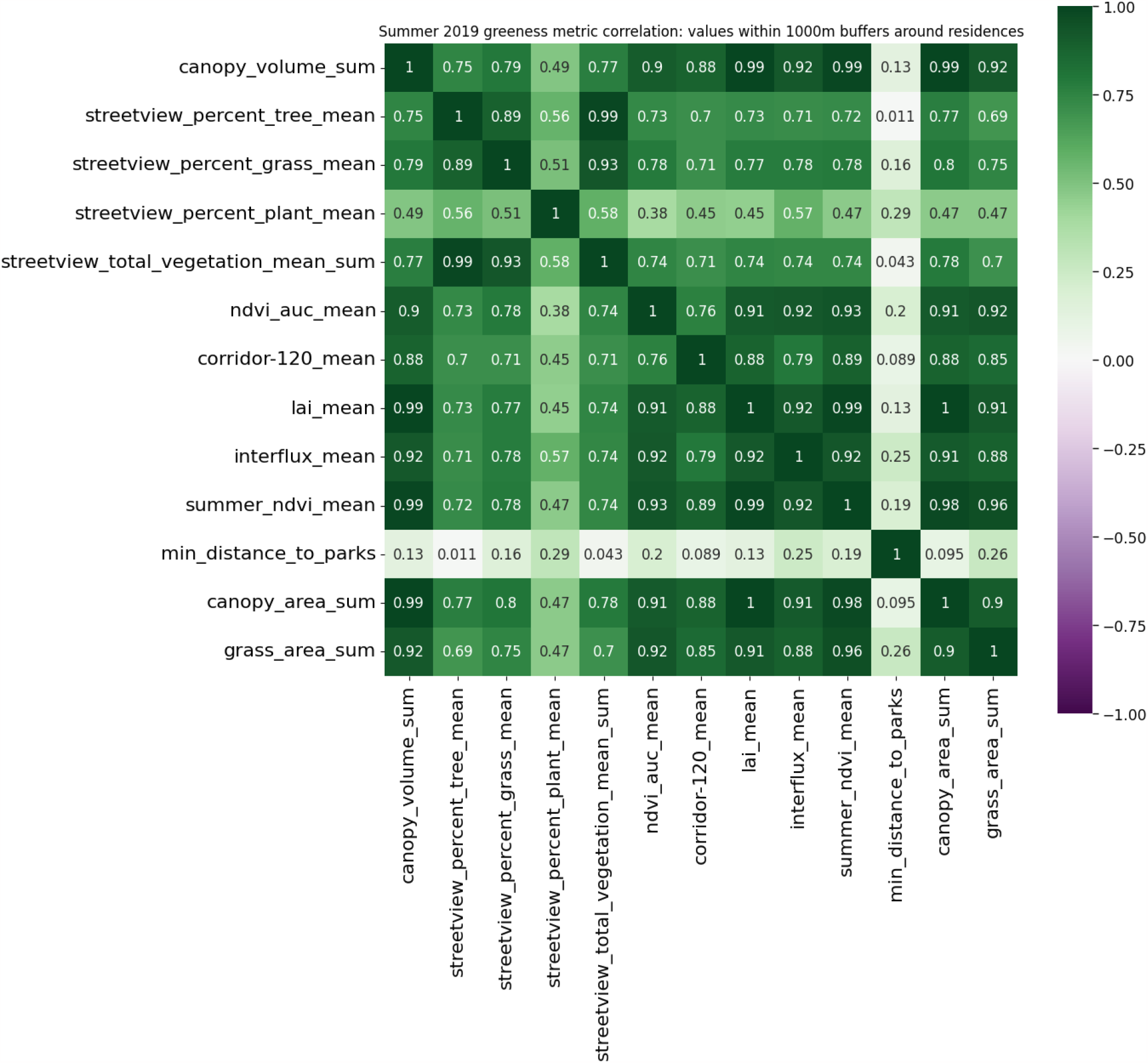
Correlation between mean metrics within a 1000 meter buffer around all address points.

Figure 6 shows the differences in trends across a few selected correlations given changes in buffer size. The pairs that are highly correlated at the 50 meter scale, such as LAI and canopy area remain so as buffer size increases, while those with lower correlations at smaller scales see larger increases with buffer size. The highly orthogonal pairs such as interaction flux and streetview, or distance to parks exhibit more of a lag with smaller increases or even small decreases between 50 and 250m, with more increasing above 250m.

**Figure 6:**
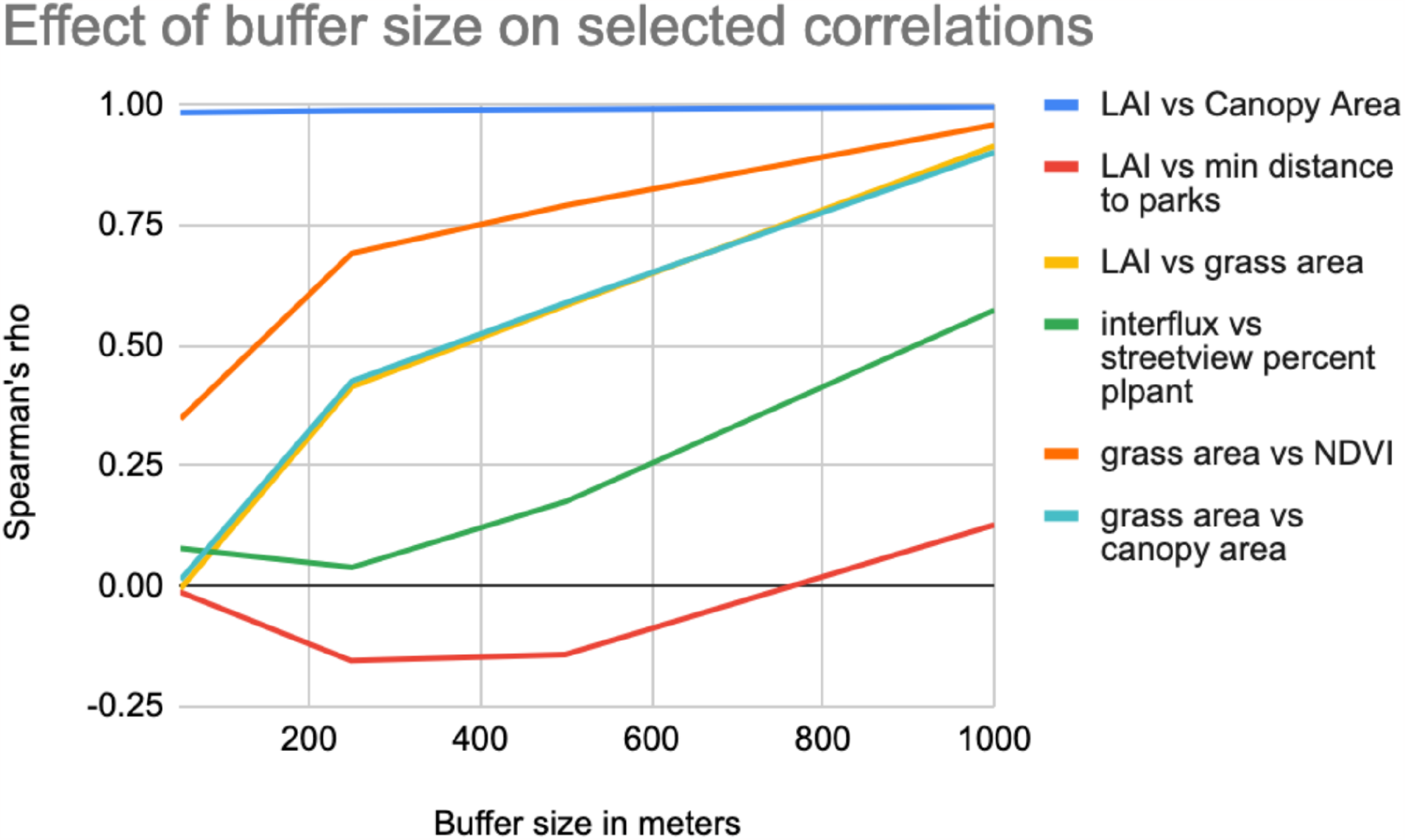
selected correlations across buffer sizes.

### PCA groupings and factor loadings

The principal components factor loadings for 50 meter buffers around addresses are shown in figure 7a. The number of principal components, 4, corresponds to that required for the explanation of around 80% of the variance. For the 50 meter buffer size the first principal component may represent overall greenness fairly well, as it has strong positive correlations with tree metrics (canopy, LAI, streetview tree), general vegetation metrics (NDVI, streetview total vegetation), and weaker but still positive correlations with grass (streetview, and to a lesser extent grass area), and the connectivity metrics associated with habitat patches (interaction flux and corridors). This component is effectively uncorrelated with minimum distance to parks.

**Figure 7:**
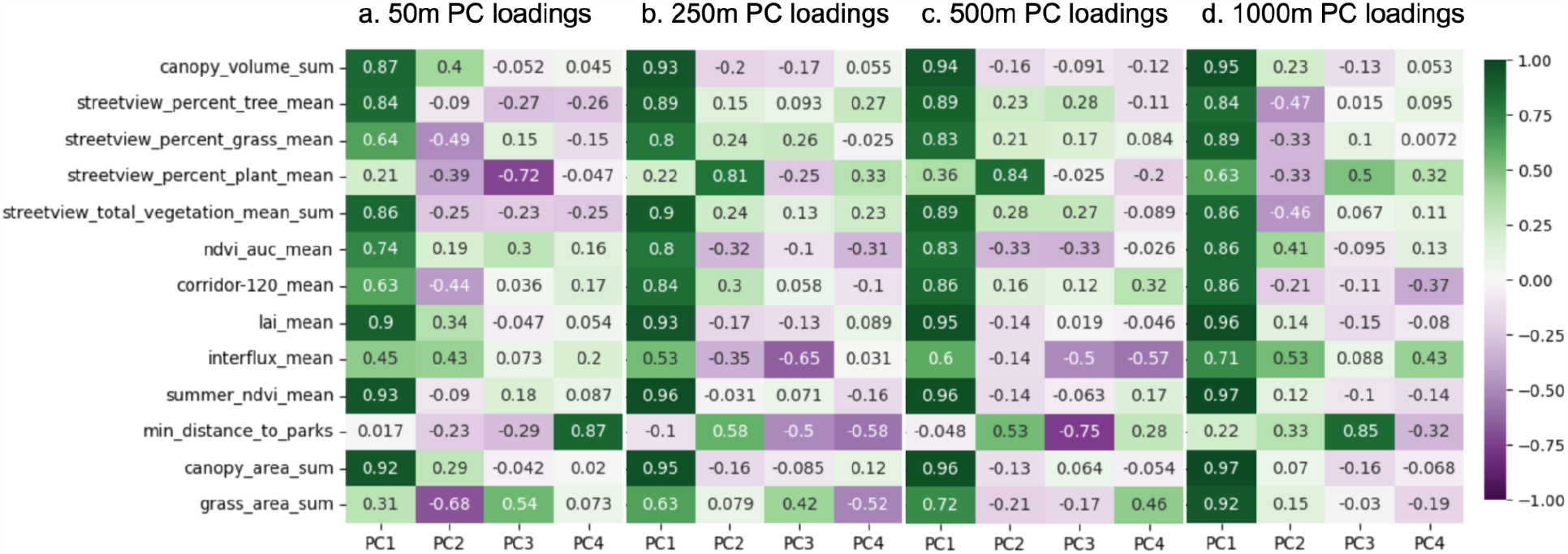
Principal component loadings for 50m (a), 250m (b), 500m (c) and 1000m (d) buffer radii.

The second principal component exhibits its strongest positive correlations with interaction flux, canopy volume and LAI, followed by canopy area and a very weak positive with NDVI AUC, but with weak negative correlations with summer NDVI, distance to parks, streetview trees, and with strong negative correlations with grass area, streetview grass and habitat corridors. This is interesting in that interaction flux, LAI and canopy volume are intuitively related to trees, but this component has no correlation with streetview trees, illustrating the fundamental orthogonality between aerial tree measurements and streetview tree measurements. Similarly this component has no correlation with summer NDVI, but shows strong correlation with LAI and tree canopy volume, illustrating the lack of tree specificity of NDVI.

The third principal component shows a strong negative correlation with streetview plant, weak negative correlations with streetview tree, corridors, LAI, distance to parks, canopy area, canopy volume, and weak positive correlations with NDVI, and a relatively strong correlation with grass area. This component seems to be representing the orthogonality of grass area and streetview plant as distinct from the other greening metrics and each other.

The fourth principal component is strongly positively correlated with minimum distance to parks and is only weakly correlated with any other metrics.

The distribution of correlations with metrics for the first principal component is similar at 250m to that of the 50m buffer data. However, in contrast to the 50 buffer, the 250m buffer second principal component does not show a strong negative correlation with grass area or streetview grass, but a strong positive correlation with streetview plant and distance to parks, while the negative correlations with LAI, canopy volume and canopy area are very weak. The third principal component meanwhile is more strongly negatively correlated with interaction flux and minimum distance to parks, with positive correlation with grass area and streetview grass. The fourth principal component has similar negative correlations with minimum distance to parks and grass area, unlike the 50m buffer which did not have this association with grass area. The principal components factor loadings for 500 meter buffers around addresses are shown in figure 7c. Here the pattern is similar to the 250m buffer, with the exception of principal component 4, which has a relatively strong correlation with interaction flux. The principal components factor loadings for 1000 meter buffers around addresses are shown in figure 7d. In this case the second principal component shows weak positive correlations with interaction flux and NDVI AUC, and to a lesser extent distance to parks, while showing negative correlations with streetview indices. Principal component 3 joins distance to parks with a streetview plant component, and the fourth principal component has weak positive correlations with interaction flux and streetview plant, with weak negative correlations with habitat corridors and distance to parks. Principal component loading plots for the 50m case are shown for component pairs 1-2, 1-3, and 1-4 in figures SI-20, SI-21, and SI-22 respectively.

### Raster only correlations

The raster-only Spearman correlation matrix is shown in figure 8. Planetscope summer average NDVI, TDVI, SAVI and GCI are all highly correlated (Spearman’s ρ of 0.98 or greater). There is a lower correlation between summer average NDVI and the NDVI annual area under the curve, which reflects the impact of seasonal phenology in the climate of the study area. We expect this difference to be less pronounced in more tropical areas and areas with a higher percentage of evergreen vegetation. In any case, for longitudinal health studies assaying health outcomes over long periods or analyzing many years of historical data, the annual area under the curve may be more appropriate as a measure of cumulative exposure to greenness than summer average NDVI or single acquisition NDVI. The ρ value between LAI and NDVI is 0.7, consistent with the intuition that these two variables are related but fundamentally reading different things. The canopy volume, canopy height maximum and canopy height minimum are highly correlated (Spearman’s ρ of 0.92 or greater). This is not surprising, as higher crown bottoms tend to manifest as trees grow taller. The least correlated variable is the corridor map, which was expected as the corridor locations as produced tend to be between rather than within the vegetation patches that confer LAI and canopy volume.

**Figure 8:**
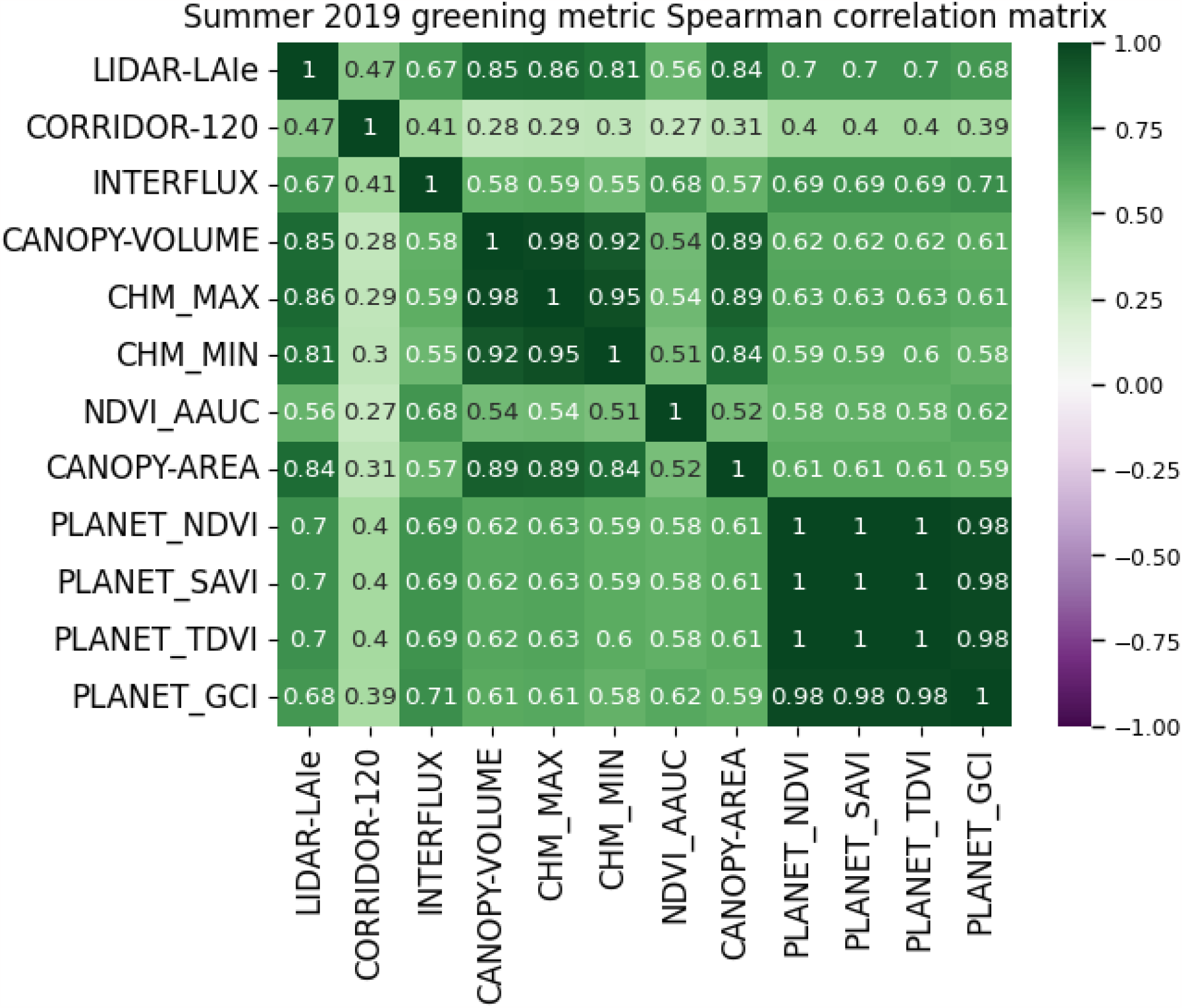
Pixel to Pixel Spearman correlation between raster datasets without residential buffers.

### Raster Regression Residuals

Figure 9 shows the linear regression residual mapping between selected raster variables. Figure 9a shows the mapping between NDVI and TDVI. As NDVI increases, TDVI saturates, so the residual map shows high values where there are edges between adjacent features. This does not suggest a categorical difference in the physical correlates of what is being measured, so much as a difference in magnitude between the metric at different values. In contrast, figure 9b shows the linear regression residual map between NDVI and LAI. The positive values show trees, negative values grass and middling values show pavement, bare earth and building roofs. While this clearly shows the difference in the physical correlates of the two metrics, with LAI showing trees, and NDVI showing trees and grass, it also draws our attention to the roads themselves, and their widths, which we can hypothesize are positively correlated with traffic counts and hence air pollution concentrations, and which are not differentiated from grass in the LAI dataset, but are likely at least somewhat proportional to the inverse of the NDVI.

**Figure 9.**
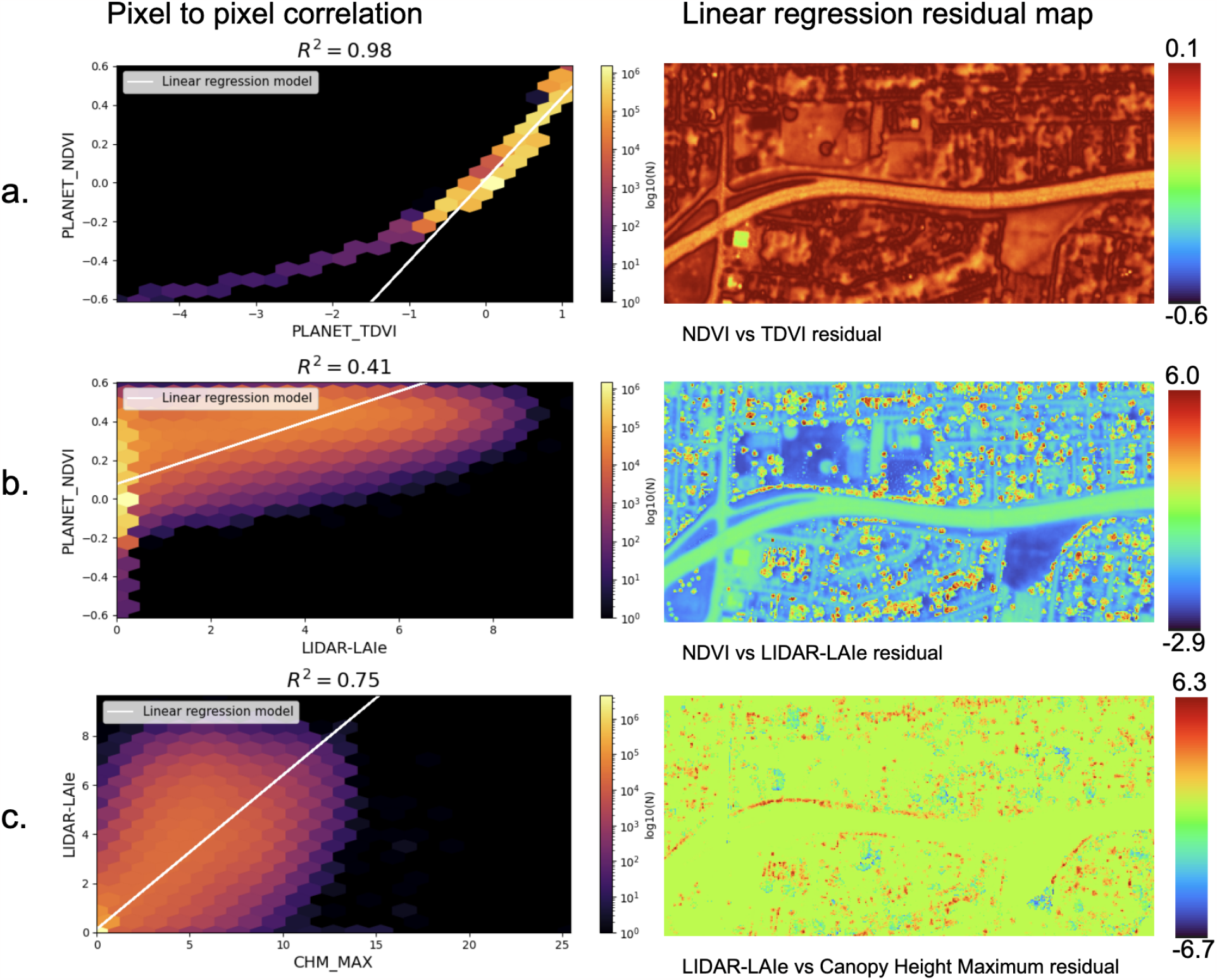
Residual maps within categories

Figure 9c shows the linear regression residual map of LAI and maximum canopy height. Here the positive values seem to represent short, dense vegetation, while the negative values represent tall, sparse vegetation. This residual map may be of interest in itself for health studies and air pollution studies, as short, dense vegetation may play a mitigating role in traffic-sourced air pollution (through deposition of particles on leaf surfaces, and dispersion of polluted air up to be diluted by cleaner air above), while tall, sparse vegetation might serve to trap traffic sourced air pollution at ground level by reducing vertical mixing. In any case it is a good illustration of the nuance required for interpreting the correlations between different greenness metrics; even though these two variables had relatively high correlation (Spearman’s ρ of 0.86), they can still clearly be associated with meaningfully different physical correlates in the environment. The residual maps shown in figure 9 are paired with their respective regressors in figures ST-15, ST-16, ST-17 and ST-18.

## Discussion

In this study, we compared 13 different greenness metrics with each other, both in the context of overall raster-based pixel to pixel correlations and also as each metric aggregated into buffers around residential addresses, to develop an understanding of the potential utility of the metrics for evaluating environmental health interventions. Of the multitude of greening metrics available to the environmental health researcher, we have found that some are more covariant than others, with many exhibiting scale-dependence in their covariation. We found the different spectral indices to be highly correlated, and so just chose NDVI to represent the group as it is most commonly used in the literature. Among the lidar derived canopy metrics, the canopy volume and max canopy height were highly correlated, which is to be expected. The bottom of canopy height was also highly correlated with these metrics, although the bottom of canopy height may be interesting in itself as it may relate to air pollution transport at the hyper-local level, in that high tree canopy bottoms may trap traffic sourced pollution at human occupied atmospheric strata, but when bottom of canopy is low enough to preclude human occupation this may become less of a health concern.

While NDVI values grass and trees similarly, leaf area index ignores grassy areas in favor of trees, which makes a bigger difference when looking at smaller buffers around addresses than at larger buffers, where the differences tend towards evening out. This effect may be different in places with different grass phenology. For example the grass stays green in Louisville all summer long, and tends to become brown in the winter, matching the phenology of the deciduous trees. However, in coastal California, the grasses turn brown in the summer, and green in the winter, out of phase with deciduous trees. Thus in places like coastal California, we can expect the correlation between summer LAI and summer NDVI to be higher than in places like Louisville, while the opposite (lower correlation in coastal California) would occur in winter.

The streetview metrics provide insights that are orthogonal to the aerial and satellite measurements. Similarly the connectivity metrics are also orthogonal, although more research is required to dial in the most relevant combination of connectivity metric parameters.

When analyzing the data aggregated into buffers around addresses, including both raster and streetview data we found that the first principal component included a broad association of greenness, including metrics that consider trees, combine trees and grass, and consider connectivity between tree patches, but not overall grass area or distance to parks. The second principal component seems tree specific, while the third principal component seems to associate with grass area, and the fourth with distance to parks. This indicates a physical interpretation of greenness, where trees and grass are distinct features that while commonly are seen together, can also be seen separately, and both trees and grass are distinct from the connectivity between them and over short distances are unrelated to the distance of an address to the nearest park.

The analysis also reinforces the orthogonality between aerial/orbial metrics and those derived from streetview cameras. The second principal component correlates with LAI, canopy area and canopy volume, but not with streetview tree index. The streetview grass index and grass area are weakly correlated. The streetview plant index is highly orthogonal to most other metrics. This latter point is of potential importance for health studies that consider the aesthetic benefits of vegetation; streetview plant index is measuring plants that are not trees nor grass, so in an urban area this means shrubs, flowers and other landscape plants that may have low total surface area but high aesthetic value, and potentially high biodiversity value.

### Implications for design

The extent to which each of the available greenness metrics is associated with community health outcomes will determine the utility of designing interventions towards changing said metrics. For example, research showing that a given increase in NDVI was associated with a particular reduction in cardiovascular disease may provide an incentive to increase NDVI of the neighborhood by that amount. From a design and engineering perspective, the most cost effective method for increasing NDVI in an urban area will depend entirely on the makeup of the built environment. In a residential urban environment dominated by single family residences, the fastest and lowest cost method of increasing NDVI might be to fertilize and irrigate lawns, and seed bare-earth areas with grass and groundcover. This is in contrast to tree planting, which requires purchasing trees, digging holes, and then waiting years for the tree canopies to increase to appreciable size. In pavement dominated areas where residents are concentrated in high-rise apartments, planting canopy forming trees may be more cost effective for increasing NDVI, as planting grass and ground covers may require expensive depaving or green-roof retrofits.

For increasing LAI, large trees, or trees that will become large will have the biggest impact, although the species of trees is very important. Needle leaf trees have higher surface area than broadleaf trees, and evergreens will have far higher annual leaf area presentation than deciduous trees. Designing for top of canopy height requires planting trees that will grow very tall. Bottom of canopy height is species dependent, if seeking a low canopy bottom, then conical conifers and hedges are appropriate, while seeking a high bottom of canopy requires tall, leggy species.

Streetview metrics that represent viewshed percentage of an object class, such as tree-view index will be most cost effectively increased by planting trees as close to roads as possible. The shape of these trees will impact their street-visible area, so wide, tall trees with low bottom of canopy may have more impact than tall thin trees or trees with tall trunks and high bottom of canopy.

Increasing interaction flux would require filling gaps between habitat patches, with a focus on linking the least connected patches to the most connected patches. There are also global connectivity measures for subregions that could be increased using the algorithm of (Clauzel et al., 2015). Decreasing distance to parks would require building new parks or enlarging existing ones. Increasing habitat corridors in the spirit of the corridor metric used in this study would require shrinking habitat patches while maintaining a minimum distance between them.

In summary, the strategy for increasing greenness in an area may be completely different depending on the metric used to define greenness. While the motivation to increase a greenness metric may be driven by the association between said metric and positive health outcomes, as more is learned about the mediators between greenness and health more mediator-specific design rubrics can be employed. For example tightly packed roadside vegetated air barriers have been demonstrated to reduce traffic-sourced pollution downwind (Al-Dabbous & Kumar, 2014), and wide canopy forming trees have been shown to decrease urban heat island effects (Schwaab et al., 2021); such interventions can be deployed, and their performance evaluated independently of any greenness metrics associated with them.

## Data Availability

All data produced in the present work are contained in the manuscript

## Acknowledgements

The authors would like to thank Chris Chandler of The Nature Conservancy for his support and encouragement in the development of novel greening metrics.

