## Supplemental Information for "Typological distinction of remotely sensed metrics of neighborhood vegetation for environmental health intervention design"

**Metrics** **2**
Spectral Indices 2
NDVI 2
Alternate Spectral Indices 3
Aerial LIDAR derived Metrics 3
LAI 3
Other LIDAR-derived canopy metrics: 4
Land Cover Classes: 4
Street view imagery: 5
Connectivity metrics: 5
**Methods:** **6**
Satellite NDVI and other spectral metrics: 6
LAI: 7
LIDAR and 18cm 4-band imagery acquisition 7
LIDAR point cloud classification 7
Leaf area index from aerial LIDAR scan 7
Canopy Volume, Canopy Height Max and Min: 8
Land Cover Classifications: 8
Streetview: 8
obtaining source images: 8
Semantic Segmentation: 9
Connectivity Metrics: 9
Distance to parks: 9
Ecological connectivity metrics: 9
**Figures:** **11**

### Metrics

#### Spectral Indices

##### NDVI

NDVI can be gathered from any imaging device that records red and infrared imagery as separate bands. Some commonly available sources for such data and their spatial and temporal resolution are summarized in table SI-1. The temporal resolution determines to what extent seasonal differences can be captured, with higher temporal resolution increasing the likelihood of finding cloud-free imagery in a given time-period. The launch date determines how far back in time data from a given source can be gathered, which is very important for longitudinal studies and studies that seek to use multi-year averages or other time-series derivatives as inputs. The spatial resolution will determine the extent to which the data reflects differences in landscape features at human scales. This is especially important for urban studies where dramatic changes in NDVI over a few meters are expected due to buildings, paved surfaces and vegetation coexisting in small spaces. While government programs such as MODIS, Landsat, Sentinel and NAIP provide data for free, commercial providers such as those operating Planetscope and Worldview satellites typically charge per square kilometer, with the price increasing with spatial resolution. For the present study we rely mainly on Planetscope data as the 3 to 4 meter resolution enables discrimination of potentially health-relevant features of urban landscapes, and the rapid return time allows consistent cloud free acquisitions for a multiple years study.

| Spectral Sensors | | | |
|---|---|---|---|
| Sensor | Spatial Resolution | Revisit Time | Launch date |
| MODIS | 250m | 1-2 days | MODIS-TERRA: 1999, MODIS-AQUA: 2002 |
| LANDSAT-8/9 | 30m | 16-8 days | LANDSAT-8: 2013, LANDSAT-9: 2021 |
| SENTINEL-2 | 10m | 5 days | 2015 |
| PLANETSCOPE | 3-4m | 1 day | 2014 |
| Worldview 3 | 0.3m | random/task | 2014 |
| Aerial USDA NAIP | 0.6-1m | 2 years | 4-band since 2007 in some states |
| For Hire, plane, helicopter or UAV | 0.03-1m | cost limited | |

Table SI-1: Common spectral data sources

##### Alternate Spectral Indices

Alternative spectral methods such as EVI (Liu & Huete, 1995), SAVI (Huete, 1988), TDVI (Bannari et al., 2002) and at least 100 other metrics (Xue & Su, 2017) can also be derived from orbital and aerial multispectral sensors. These have been developed to overcome some of the limitations of NDVI with respect to atmospheric effects, soil composition differences, vegetation composition and other factors. For the present study we have calculated the TDVI, SAVI, SIPI and GCI.

#### Aerial LIDAR derived Metrics

##### LAI

**Leaf area index:**

Leaf area index (LAI) is a biophysical parameter of ecosystems that expresses vegetation foliar surface area in units of leaf area per unit of ground surface area ((Watson, 1947)). LAI is related to many important ecological functions of vegetation, including photosynthetic primary production (Gitelson et al., 2014), carbon cycles (Q. Li et al., 2018) and biodiversity (Peng et al., 2016).  LAI is also related to evapotranspiration (Yan et al., 2012), which along with shade drives urban heat island effect mitigation (Paschalis et al., 2021), (Hardin & Jensen, 2007). Furthermore, LAI is related to the deposition and dispersion of air pollutants by urban vegetation (Janhäll, 2015), which may make it a key variable for understanding the human health benefits of urban vegetation.  A few health studies have adopted LAI as a greenness exposure metric (Orioli et al., 2019), which has shown associations with health outcomes in studies where tree canopy area did not (Jennings et al., 2019), indicating the surface area specificity of LAI may have predictive consequences for health studies over and above LAI's proportionality to canopy area. However, the limited availability of high spatial and temporal resolution urban LAI data has become an obstacle for many environmental health researchers.

LAI measurements are broadly categorized as using direct and indirect methods (Jonckheere et al., 2004). Direct methods, such as collecting leaf-fall or denuding portions of trees, followed by measurement of each leaf's size (Sellin, 2000), or weighing the leaves and applying a conversion factor (Bréda, 2003) are not practical at the spatial scales required for health studies or responsive environmental interventions. Appropriately scaled indirect methods utilize remote sensing platforms to estimate the LAI. These methods can be casually grouped into spectral methods and geometric methods. Spectral methods infer the LAI from multi or hyper-spectral sensor measurements via regression analysis (Lee et al., 2004), usually ground-truthed with geometric measurements such as hemispheric photometry (Propastin & Erasmi, 2010). Spectral methods are limited to the spatial and temporal resolution of the chosen spectral sensor sensor. For example, LAI from MODIS satellites (Wenze et al., 2006) is gathered with 1km pixels at 8 day revisit time, Landsat 8 derived LAI (Blinn et al., 2019) provides estimates at 30m resolution every 16 days, while Sentinel-2 derived LAI (Hu et al., 2020) provides 10m resolution every 5 days. The regression equations used to estimate the LAI from the spectral information must be generated for specific biomes, as the relationship between LAI and spectral signatures varies for different plant communities and species. While high correlation coefficients are seen in non urbanized environments such as rice paddies (Wang et al., 2011), wheat fields (X. Li et al., 2014), Temperate deciduous forests (Meyer et al., 2019) and Alluvial forests (Tillack et al., 2014), urban areas present a complex problem for this approach, as the spatial distribution of vegetation communities and other spectral features is highly heterogeneous, and it is these diverse spatial elements that are the subject of greening interventions for improving community health.

Geometric methods of estimating LAI from aerial LIDAR data overcome some of the limitations of spectral methods employing satellite imagery for urban areas. LAI can be estimated from aerial LIDAR using variations of the Beer-Lambert law (Richardson et al., 2009), which takes into account 3 dimensional point density sequences to relate the canopy absorbance of laser pulses with LAI. This is similar to (but inverted from) hemispheric photometry and leaf canopy analyzer methods (Bréda, 2003) which also employ Beer's law by measuring the obstruction of skylight by leaves. Using Beer's law, LIDAR based LAI estimates can be obtained for an area with spatial resolutions smaller than 1m, which are responsive to varying phenological conditions. While such geometric methods are potentially far more precise for urban LAI estimation than spectral methods, the availability of aerial LIDAR point clouds for many areas is quite limited, although this is changing rapidly. Aerial LIDAR data is often gathered by governments seeking to make precision topographic maps, with some US states flying LIDAR every few years. These datasets are typically either flown in winter (to obtain terrain information without obstruction from deciduous canopy cover) or are flown over the course of multiple years in all seasons, leading to seasonal differences in deciduous leaf cover throughout the scan area. For the present study, we rely on a summer of 2019 aerial LIDAR scan to produce a 1m resolution LAI map.

##### Other LIDAR-derived canopy metrics:

Aerial LIDAR scans can also be leveraged to produce estimates of tree canopy volume and height, and these metrics can be associated with health metrics. For example (Chi et al., 2022) found that higher canopy volume was associated with decreased medication sales. Canopy volume can be calculated from LIDAR data in several ways. (Chi et al., 2022) segmented trees from the top of canopy height raster using a watershed algorithm, and then took the convex hull of the tree crown points to create crown volumes. Because our study area has many deciduous trees of complex crown shapes, a watershed algorithm would not work for segmenting our trees. Thus we instead divided the top-of-canopy height map and the bottom-of-canopy height map into 1 meter grids, and took the difference between them as the volume in cubic meters of canopy volume per meter squared of land area.

#### Land Cover Classes:

Land cover classes such as tree canopy, building footprints, grass, bare earth and pavement are segmented from aerial or satellite imagery. At the spatial resolution required for differentiating urban features (1 meter) these are made by combining aerial LIDAR data with multispectral information to infer the different feature classes based on color, height, and LIDAR point cloud attributes such as linearity and planarity. Tree canopy area derived from land cover class datasets is often used as a greenness metric for health research.

#### Street view imagery:

Freely or inexpensively available street-view photographic images are generated by Google for many areas, with similar products available from Mapillary, Tencent, Baidu, Bing, and others. These have been leveraged for ecological analysis; for example, *Green View Index* (GVI) refers to a method to quantify the street level amount of greenery in the view of Google Street View (GSV) Cameras. As such the GVI is a measure of street-accessible greenness of viewsheds. GVI is calculated by counting green pixels in GSV panoramas, and hence represents street level human perceptible greenness (X. Li et al., 2015). GVI has been found to have a negative correlation with certain disease measures (X. Li & Ghosh, 2018, Leng et al., 2020), and composite indices employing green-view and NDVI been associated with socioeconomic variables (Larkin & Hystad, 2019). However, counting green pixels does not differentiate between trees, grass and other non vegetative green objects, and hence provides limited actionable guidance for design decisions. Existing geometric class segmentation algorithms can be used to classify trees and grass from GSV images (Seiferling et al., 2017). Deep learning algorithms have also been used to classify trees (Li 2021) and other air-quality related built environment features of street-view images (Hu et al. 2020). Deep learning derived streetview greenery has been associated with health outcomes (Helbich et al. 2019), and this association has been found for health measures that are not associated with NDVI (Wang et al. 2019). For the present study we gathered Google street-view imagery from the summer of 2019 and used a semantic segmentation algorithm called PSPnet (Zhao et al. 2016), trained on the ADE20k dataset (Zhou et al. 2016). The ADE20k dataset segments trees, grass, plants, flowers and palms as part of 150 semantic categories, enabling fine tuning of the greening metric variables understudy. PSPnet was also used by (Qi & Hankey, 2021) to use street view semantic viewshed analytes as predictors for hyperlocal air pollution regression.

#### Connectivity metrics:

Distance to parks is the minimum distance (Euclidean or Rectilinear) between an address and the closest park. This metric has been associated with physical activity and mental health outcomes. The metric can be expanded to a distance to greenspace generally. The metric could also be modified to account for the impact of transit options, traffic conditions and path quality to make a greenspace accessibility index. While distance to parks relates to the connectivity between human residences and greenspace, there also ecological connectivity metrics that reflect the connectivity between habitat patches for all species. While perhaps straying away from metrics of "greenness" per se, these ecological connectivity metrics may also be important for understanding the health effects of the built environment, and it is important to understand how these may be more or less co-variant with other greenness metrics. For example plant biodiversity has been linked to a decreased risk of asthma, where overall greenness measured by NDVI increased risk, when controlling for air pollution as a covariate (Donovan, Landry, & Gatziolis, 2021). Plant biodiversity was strongly correlated with decreased childhood leukemia (Donovan et al. 2021), with a hypothesized mechanism involving immune system regulation via the biodiversity hypothesis of inflammatory disease, which posits that decreased microbiome diversity contributes to chronic inflammatory and autoimmune disease through undereducation of the immune system (Haahtela et al. 2013). Leaf surface area metrics such as LAI intuitively relate to such hypotheses by providing the substrate for the foliar microbiome (Stone et al. 2018), and indeed leaf area has been correlated with biodiversity in general (Peng et al. 2017). Other structural properties of vegetation (vertical stratification and complexity), have been shown to strongly correlate with nearby airborne microbial biodiversity (Robinson et al. 2021). Indeed, diversity of street-tree species has recently been shown to correlate with reduced risk of cardiovascular and stroke mortality (Giacinto et al. 2021).

Direct measurement of biodiversity can be a costly and time-consuming process. However there are ecological metrics that are more amenable to remote sensing methods at large scale. For example there are myriad connectivity metrics that can be derived from landscape features (Kindlemann & Burel, 2008), though the choice of which to use and in what configuration for a particular situation is complicated by the large number of available metrics and parameters that define them (Keeley, Beier, Jenness, 2021). From an intervention design perspective, connectivity is of particular interest as it creates a large scale of value for tree planting locations, where a single well-placed tree might have a higher impact than 100 trees planted in a less critical location. It is beyond the scope of the present study to exhaustively weigh the applicability of all possible connectivity metrics and their configuring parameters, however by way of providing some example datasets to include in the comparison with other greenness metrics we have generated two connectivity metrics for the study area. The first is interaction flux first described by Foltête, Girardet & Clauzel (2014), which is a value assigned to landscape elements (for example habitat patches, in our case tree canopy patches) that corresponds to each element's contribution to the probability of connectivity (PC), described by Saura & Pascual-Hortal (2007). The second is a spatial count of potential habitat corridors connecting patches given a maximum distance. Both of these metrics were calculated using Graphab software (Foltête et al. 2021).

### Methods:

#### Satellite NDVI and other spectral metrics:

3 to 4 meter spatial resolution 4 band satellite images were selected from cloud free days and purchased from Planet Lab's Planetscope archive. All image manipulation was performed using the Python 3.8.10 Rasterio library version 1.2.10. The Planetscope constellation uses multiple generations of small satellite technology, and so images acquired from the Planetscope 0 (dove-classic / dove / PS2) were rectified with the Planetscope 1 (dove-R / PS2.SD) using the correlation coefficient established by Huang & Roy (2021). The following spectral greenness metrics were then calculated for each image acquisition:

$$NDVI = \frac{NIR - Red}{NIR + Red}$$

$$TDVI = 1.5\frac{NIR - Red}{\sqrt{NIR^2 + Red + 0.5}}$$

$$SAVI = \frac{1.5(NIR - Red)}{NIR + Red + 0.5}$$

$$GCI = \frac{NIR}{Green} - 1$$

We averaged the metrics on 2019/05/17, 2019/06/03, 2019/06/21. 2019/07/13, 2019/08/16, 2019/09/12 and 2019/09/22 to make a mean summer 2019 dataset for each metric. The summer average NDVI, TDVI, SAVI and GCI are shown in figures SI-2, SI-3, SI-4, and SI-5 respectively. The above dates were combined with acquisitions from 2019/01/05, 2019/01/25, 2019/02/16, 2019/03/05, 2019/03/16, 2019/10/05, 2019/11/01, 2019/11/10, 2019/12/11, 2019/12/23 to create full year time series for NDVI. We used the Python Numpy library version

1.22.3 to integrate each pixel of the annual time series using the composite Simpson's rule to yield a single raster representing the annual area under the curve for NDVI values, shown in figure SI-6.

### LAI:

#### LIDAR and 18cm 4-band imagery acquisition

LIDAR data was gathered from August 15, 2019 to August 17, 2019 in 3 flights by Quantum Spatial Inc. (Lexington, KY) using a Leica ALS70 LIDAR sensor flown at an altitude between 1167 and 1270 meters, at a ground-speed of 120 kts. The sensor was set at a scan rate of 69.5 Hz, and a laser pulse rate of 221.1 kHz. With a 15° field of view and 334m wide swaths spaced 150.3m apart, the resulting point cloud density averaged 19.77 points per $m^2$.

4-band imagery was acquired by Quantum Spatial on September 9th, 2019 in a single flight at 12500 ft of altitude at a ground speed of 150 kts, using an UltraCam Eagle RGB-NIR sensor with 100mm focal length. Imagery swaths were 16400 ft wide with overlap of 55%, yielding a ground sampling resolution of 0.6 ft.

#### LIDAR point cloud classification

The LIDAR data acquisition resulted in 216 tiles of 301000$m^2$ each. Each LIDAR point cloud tile has a corresponding 4-band, 18cm raster tile. The 4-band tiles were processed into NDVI tiles, and the NDVI tiles used to colorize the LIDAR point cloud tiles using Point Data Abstraction Library (PDAL 2018) so that each point in the LIDAR point cloud has an NDVI value. The NDVI tiles were studied to determine a single NDVI threshold value to distinguish between vegetation and non vegetation (which in this case was 0.23). Ground points and above-ground points in the point cloud were then separated using the cloth simulation filter of Zhang et al. (2016). Rooftops were then separated from the off-ground points using the PDAL coplanarity filter (based on Limberger & Oliveira 2015), and then added back to the ground points to create a below canopy layer. The remaining off-ground points were filtered by the NDVI threshold to yield a canopy layer of off-ground, non-planar points with NDVI above 0.23.

#### Leaf area index from aerial LIDAR scan

The estimation of LAI from aerial LIDAR scans (ALS) has been approached in several ways; for a comprehensive review see Wang & Fang (2020). The approach that we use employs the Beer-Lambert law as proposed by Solberg et al. (2006). This method was compared to other methods by Richardson et al. (2009) and found to produce the best results. Klingberg et al. (2017) further validated the method against ground-truth in an urban environment. Using the Python 3.8.10 Numpy library version 1.22.3, the points classified as canopy and below-canopy were separately subdivided into 3ft by 3ft horizontal grids, resulting in a set of pairs (canopy and below-canopy) of 3$ft^2$ cells. To each pair of cells the following equation was used to obtain the estimate of the LAI (LAIe):

$$LAIe = -\left(\frac{cos(\theta)lidar}{0.5}\right) \times ln\left(\frac{below\ canopy\ \ returns}{below\ canopy + canopy\ returns}\right)$$

Where $(\theta)_{lidar}$ is the average scanning angle of the LIDAR sensor for the grid-cell. The resulting 3$ft^2$ LAI estimates were rasterized and resampled to 1$m^2$ by averaging the values to a 1 meter grid, resulting in the georeferenced 1$m^2$ LAI estimates shown in figure SI-7.

##### Canopy Volume, Canopy Height Max and Min:

Canopy volume was calculated from the vegetation points and ground points segmented during the LAI process discussed above. Using CloudCompare (CloudCompare, 2022) the ground points were subsampled to octree level 6, and then used to make a mesh via DeLauney triangulation with a maximum edge length of 100 meters. Then the canopy points were loaded and assigned point-to-mesh distances to the nearest edge of the ground mesh. The canopy points were then rasterized at a resolution of 1 meter using the maximum and minimum point-to-mesh distances to get the top of canopy and bottom of canopy rasters respectively. The difference between these was the canopy volume index (CVI) in units of cubic meters per square meter of ground area. The sum of the CVI within any given areal buffer becomes the total canopy volume within that buffer. The canopy volume index, canopy height maximum and canopy height minimum maps are shown in figures SI-8, SI-9 and SI-10 respectively.

#### Land Cover Classifications:

We downloaded 1.5 meter resolution 7 class land cover representing 2019 conditions from the Louisville and Jefferson County, KY Information Consortium open geospatial data website. This dataset was created for the Louisville urban tree canopy study by the University of Vermont spatial analysis laboratory (O'Neil-Dunne & Safavi, 2021). The dataset includes a raster with 7 classes: roads, grass, trees, parking lots & driveways, building footprints, bare earth and

#### Streetview:

##### Obtaining source images:

Panoramic streetview images were downloaded from Google using the street view API. Road centerlines for the neighborhood were obtained from Open Street Map using the Overpass API (http://overpass-api.de/), and points placed every 20 meters along each roadline. This resulted in 13,406 points. For each point, Google's streetview api was queried for the nearest panorama, and the 90 degree x 90 degree square images downloaded for each of the north, south, east, west and up portions of the spherical panoramas.

##### Semantic Segmentation:

Each downloaded streetview image is processed with PSPnet trained on ADE20k, to yield categorization of each pixel in the image within any of 150 semantic categories in the ADE20k dataset. For each location the total pixel count of each category for each compass direction and up were summed to get the total pixel count for each location. This was divided by the total pixels per location to get the viewshed index for each category per location. Of the 150 ADE20k categories, for this study we focused on the Tree view (shown in figure SI-11), grass view, total vegetation view (sum of tree, grass, plant, flower, and palm).

#### Connectivity Metrics:

##### Distance to parks:

GIS files for the addresses for every building, and the polygons of every park in the study area and were obtained from the Louisville and Jefferson County, KY Information Consortium open geospatial data website. We used the Python 3 geopandas library's distance function to calculate the minimum distance between each address point and the nearest park. Figure SI-19 shows the address points colorized by distance to the nearest park.

##### Ecological connectivity metrics:

A total canopy polygon layer was generated in Qgis 3.24.3-Tisler by creating a binary mask of the LAI raster where every pixel with an LAI greater than 0 was assigned a value of 1, and all other pixels assigned a value of 0. A polygon was then drawn around the pixels with value of 1 using the Qgis polygonize tool. The resulting canopy polygons were loaded into Graphab software (Foltête et al. 2021) as a habitat patch layer. A weights layer was made in Qgis by inverting the LAI data using the Qgis raster calculator as follows:

$weight \ =- \ 1\, LAI \ + \ LAI_{max}$

The Graphab linkset generator made a linkset from the canopy polygons using the weight layer as weights. To generate the corridors the maximum distance variable was set to 120. The max weight in the weight layer of around 12 corresponds to LAI of 0, so a maximum distance of 120 corresponds to roughly 10 meters of unvegetated distance. The resulting corridor map is shown in figure SI-12. This variable can be adjusted to consider specific types of corridors, for example some birds might have a much larger max distance, while some microorganisms might have a much smaller one.

The same linkset was used to generate the interaction flux map. Interaction flux is the local contribution of each habitat patch to the probability of connectivity:

$$IF_i = \sum_{j=1}^{n} a_i^{\beta} a_j^{\beta} e^{-\alpha d_{ij}}$$

Where $IF_i$ is the interaction flux for habitat patch $i$. The area of a patch is denoted by $a$, β is a weight capacity coefficient and $e^{-\alpha d_{ij}}$ is the probability of movement between patches $i$ and $j$.

To generate the interaction flux map, parameters $d$ and β were set at 10 and 0.001 respectively. The resulting interaction flux map is shown in figure SI-13.

### Figures:

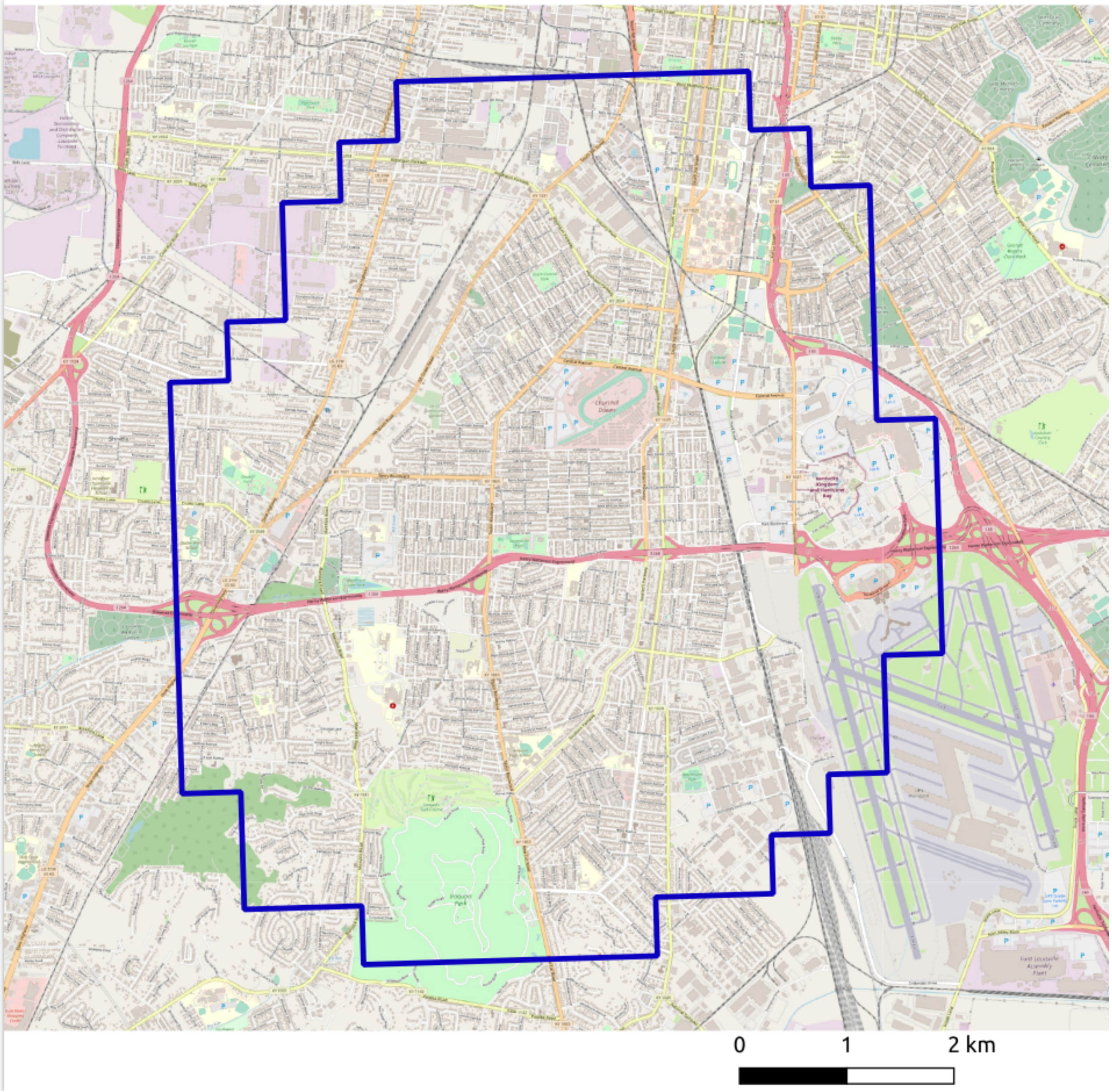


*Figure SI-1: The extended study area used for raster-to-raster analysis*

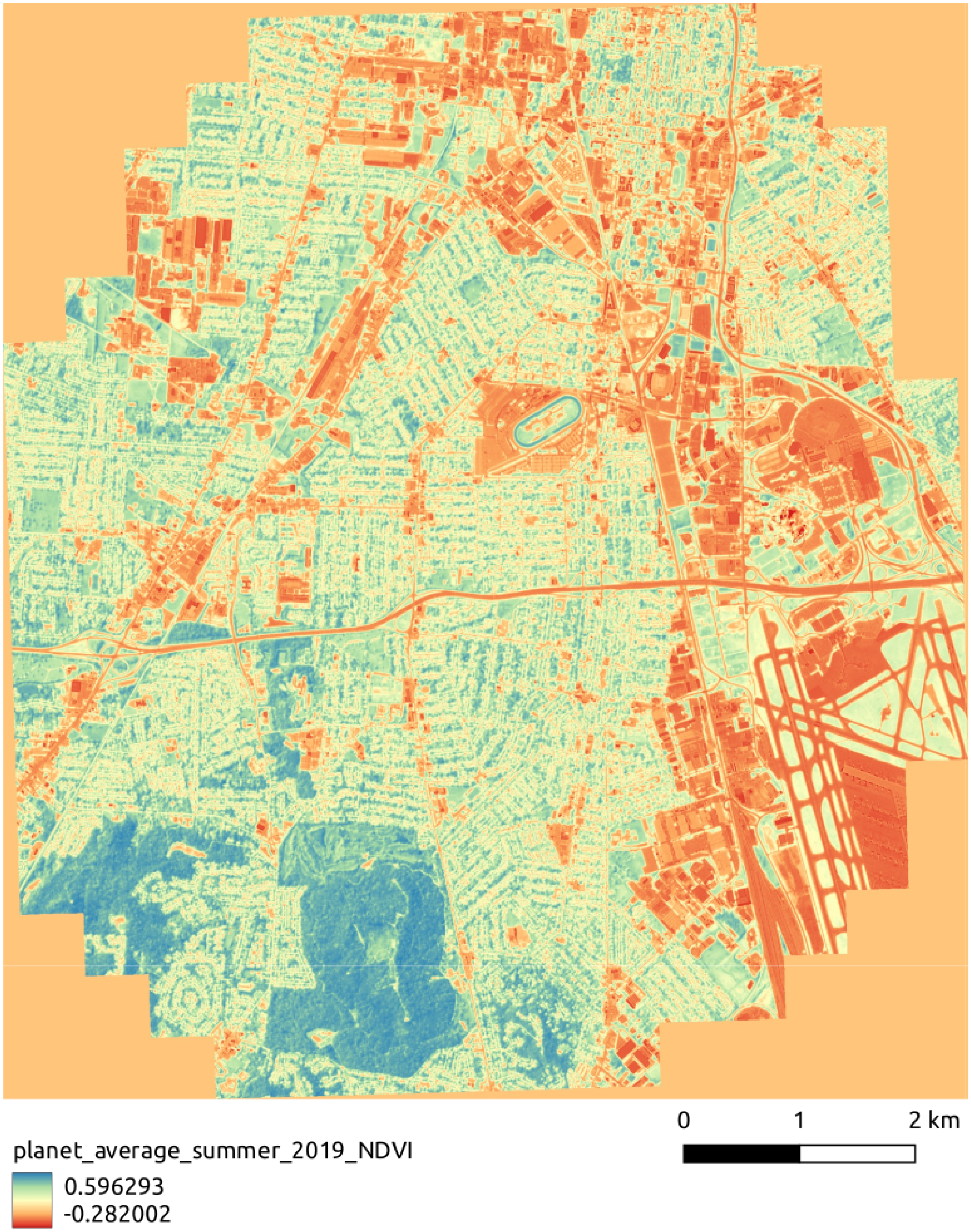


*Figure SI-2. The 4 meter average summer 2019 NDVI*

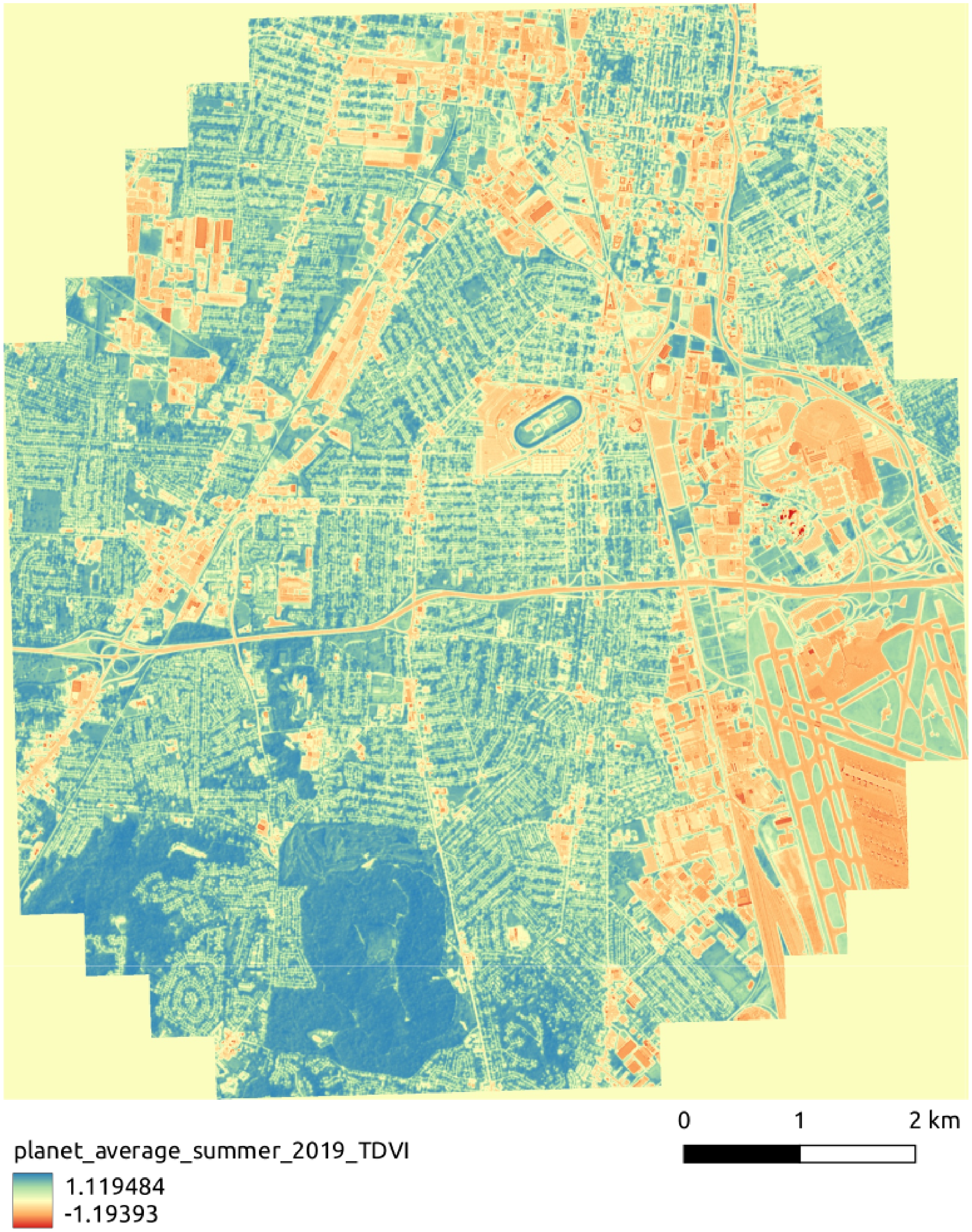


*Figure SI-3: The summer 2019 average 4 meter TDVI*

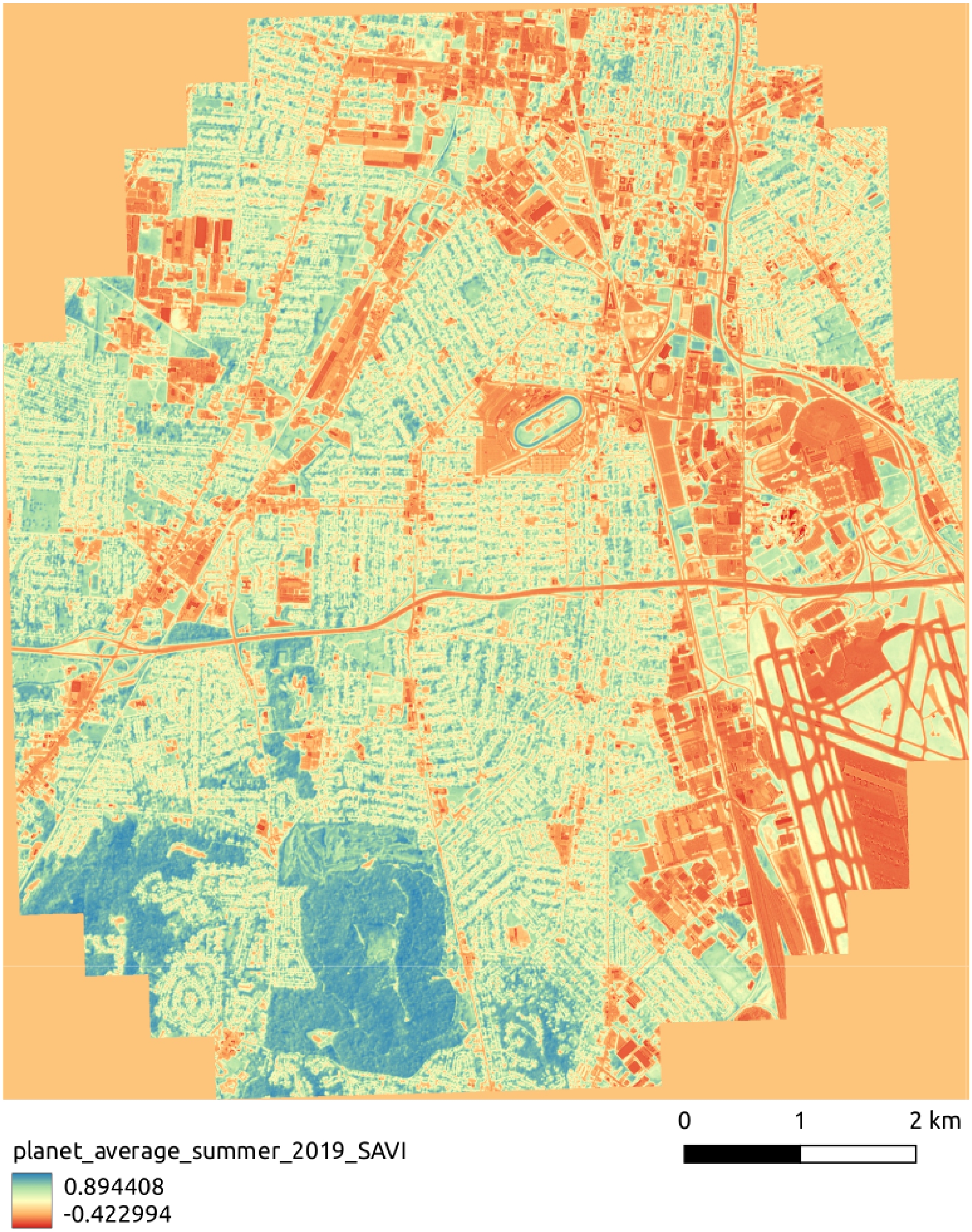


*Figure SI-4: The average summer 2019 4 meter SAVI*

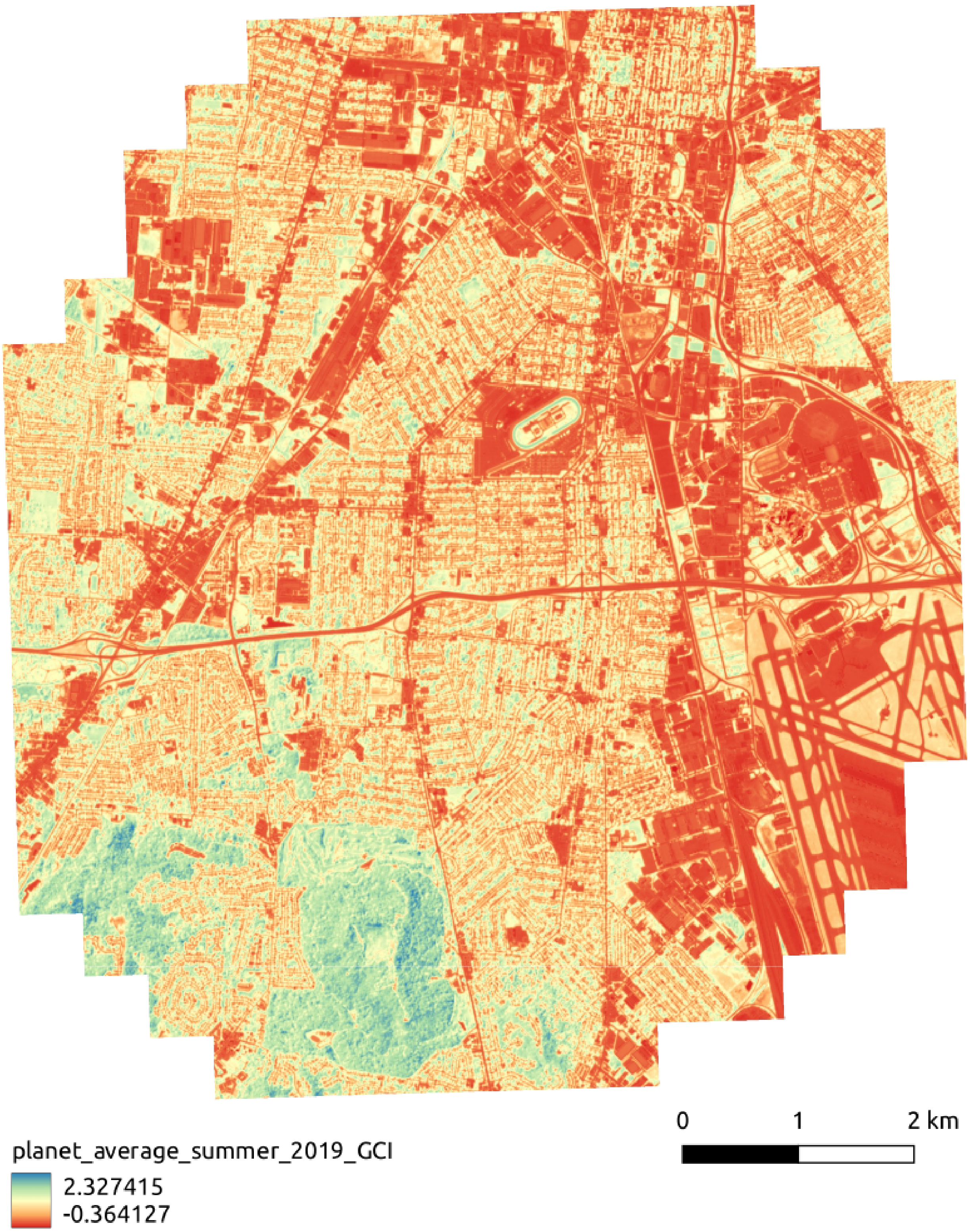

*Figure SI-5: The average summer 2019 4 meter GCI*

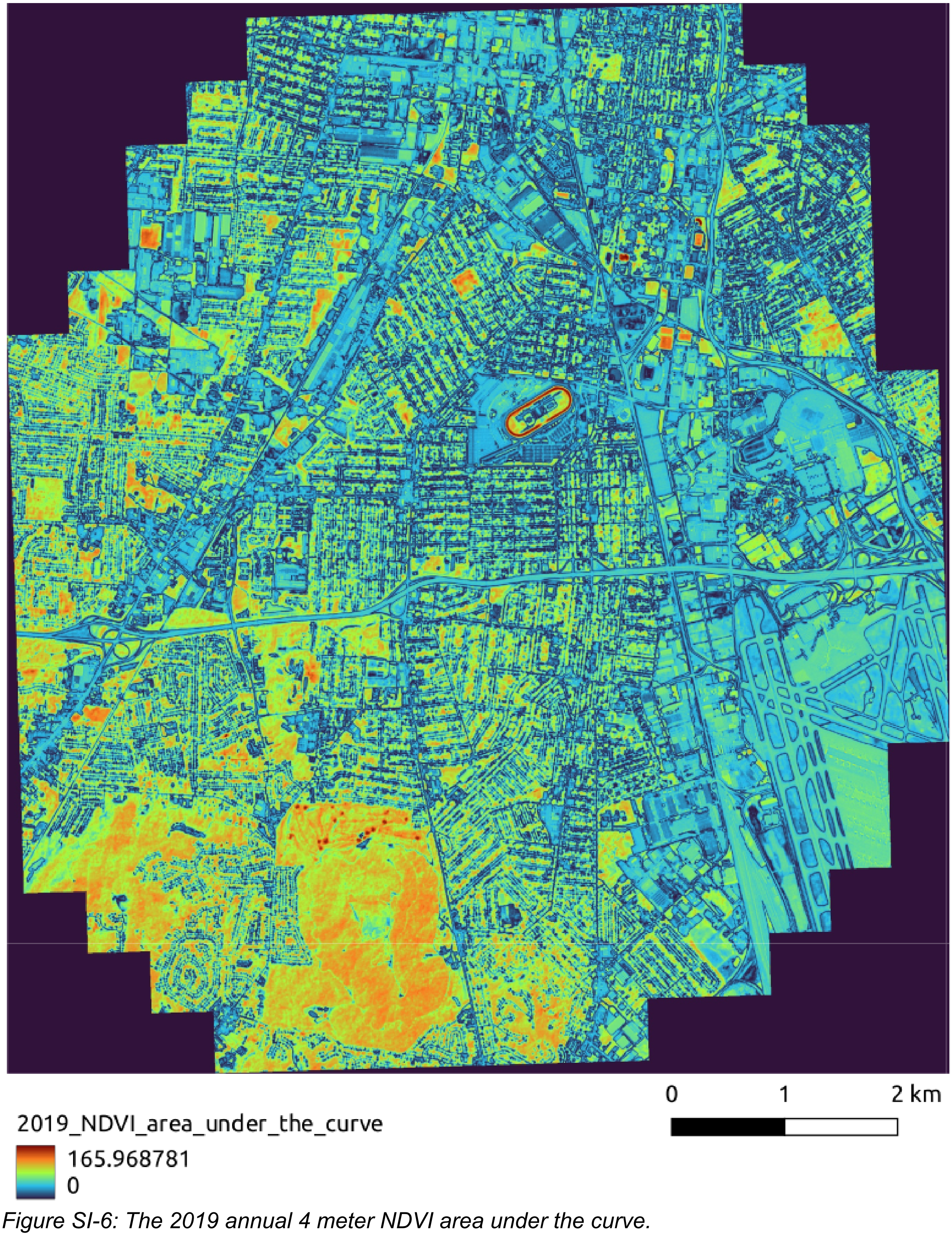


*Figure SI-6: The 2019 annual 4 meter NDVI area under the curve.*

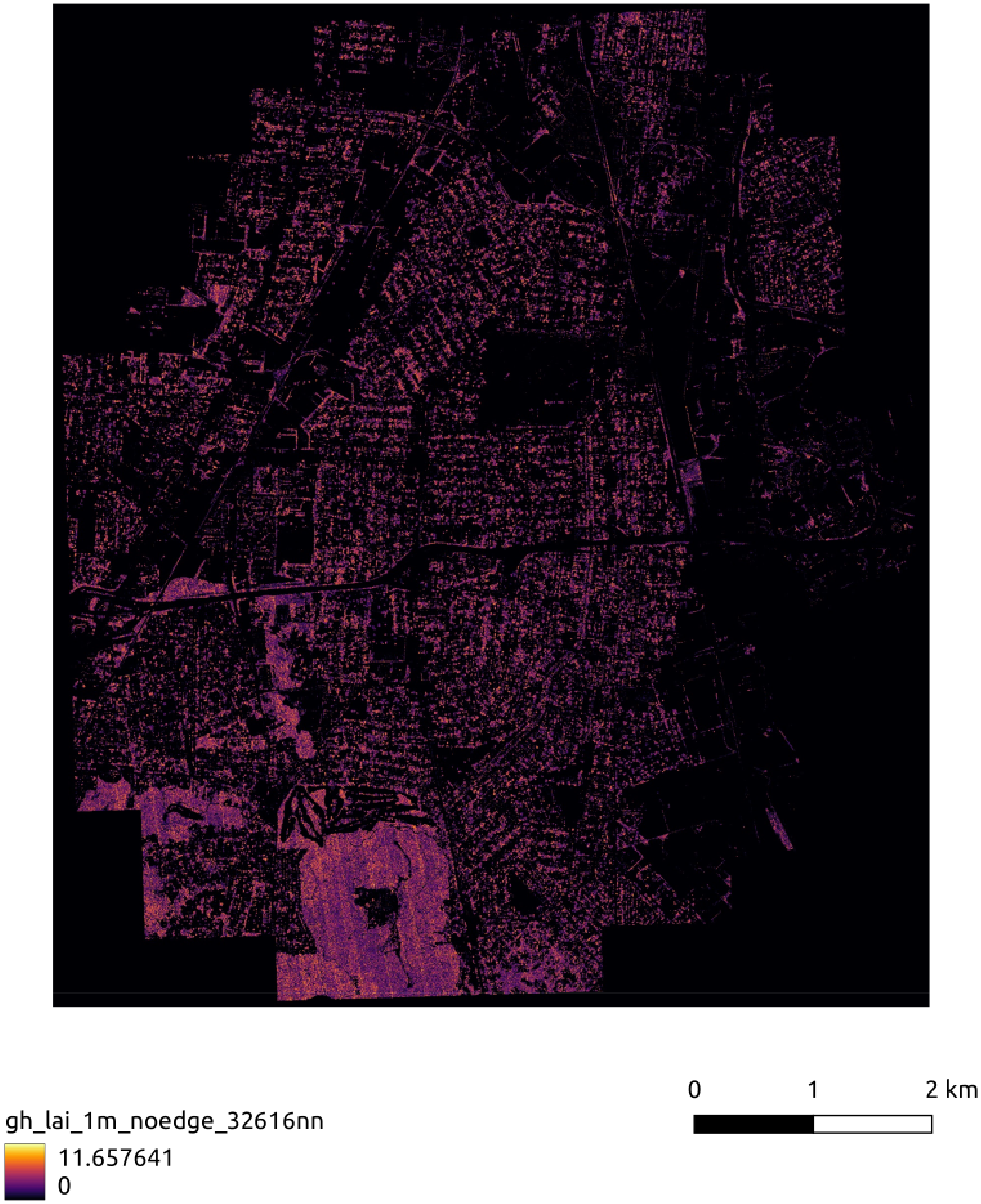


*Figure SI-7: The 1 meter resolution Leaf Area Index (LAI) estimated from aerial LIDAR data.*

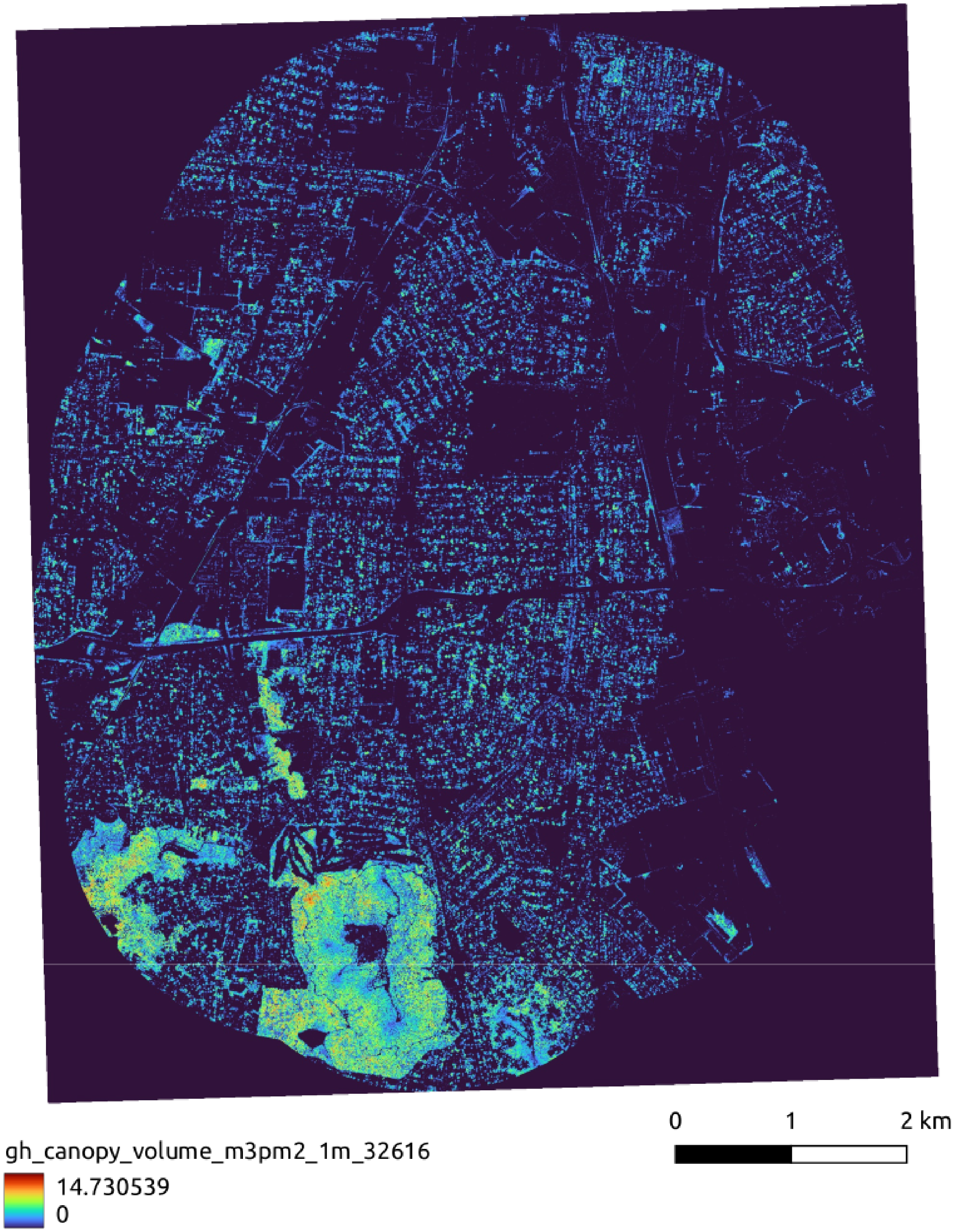


*Figure SI-8: Canopy Volume Index in cubic meters per square meter, derived from aerial LIDAR.*

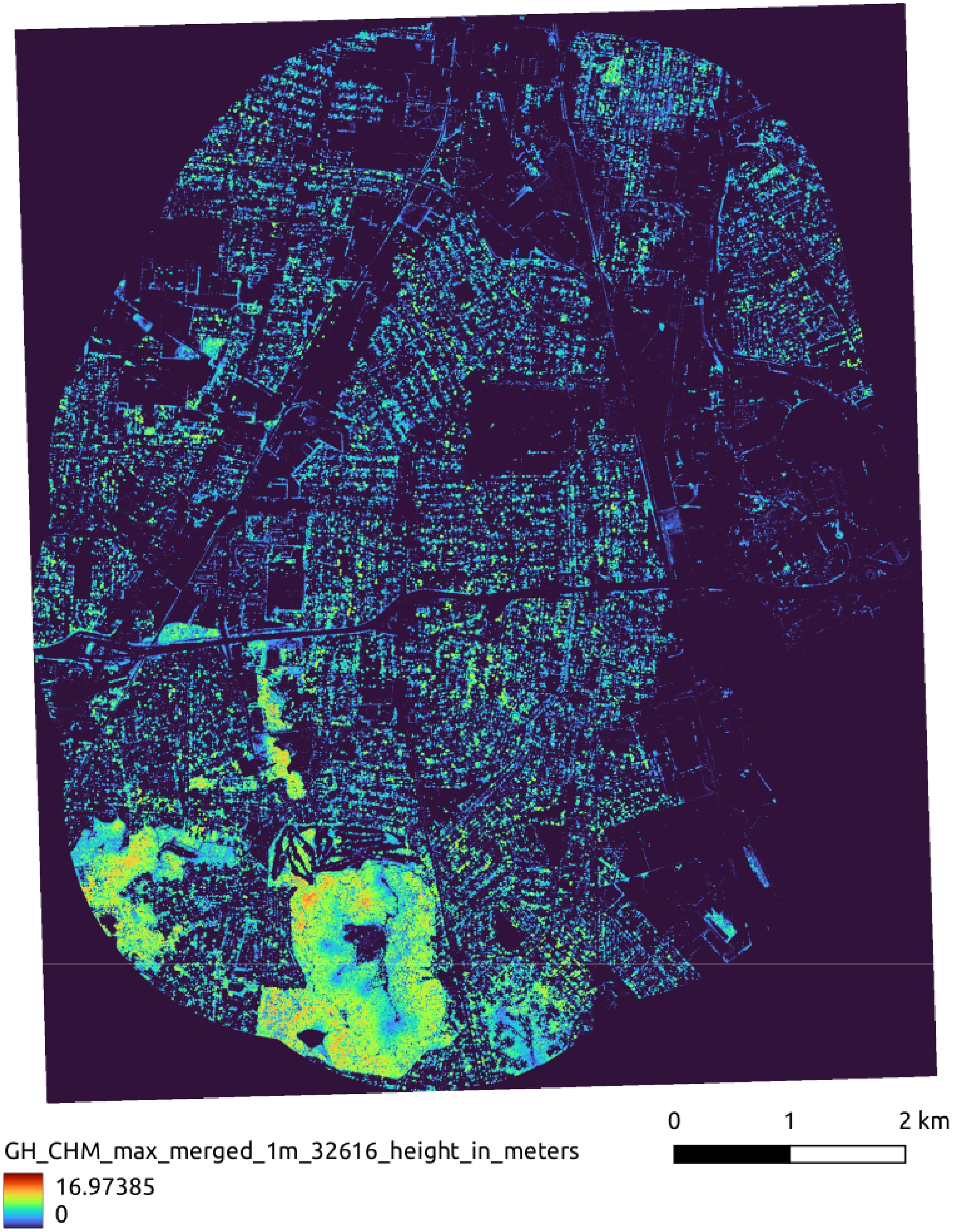


*Figure SI-9: Maximum canopy height in meters, at 1 meter resolution derived from aerial LIDAR.*

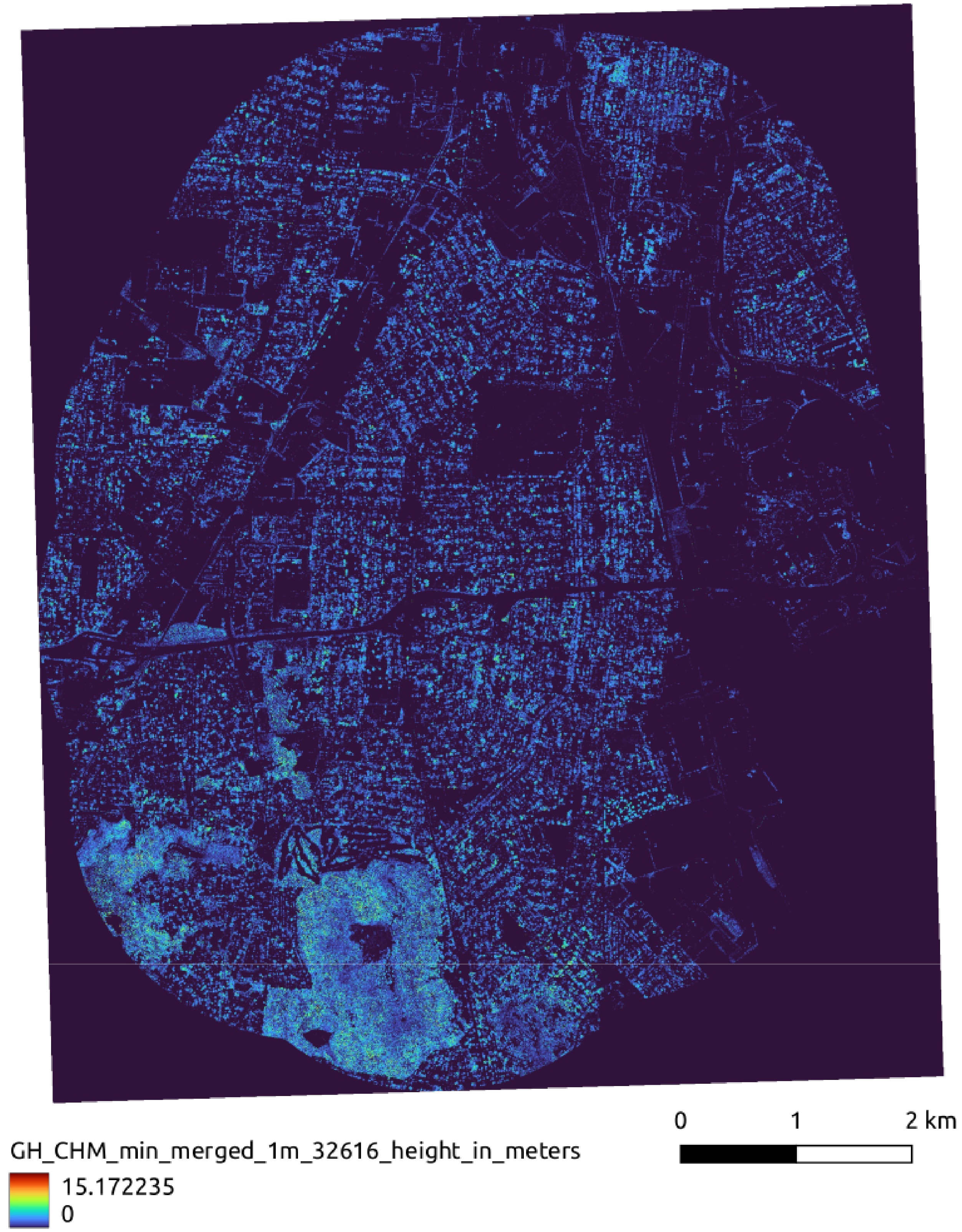


*Figure SI-10: Bottom of canopy height in meters.*

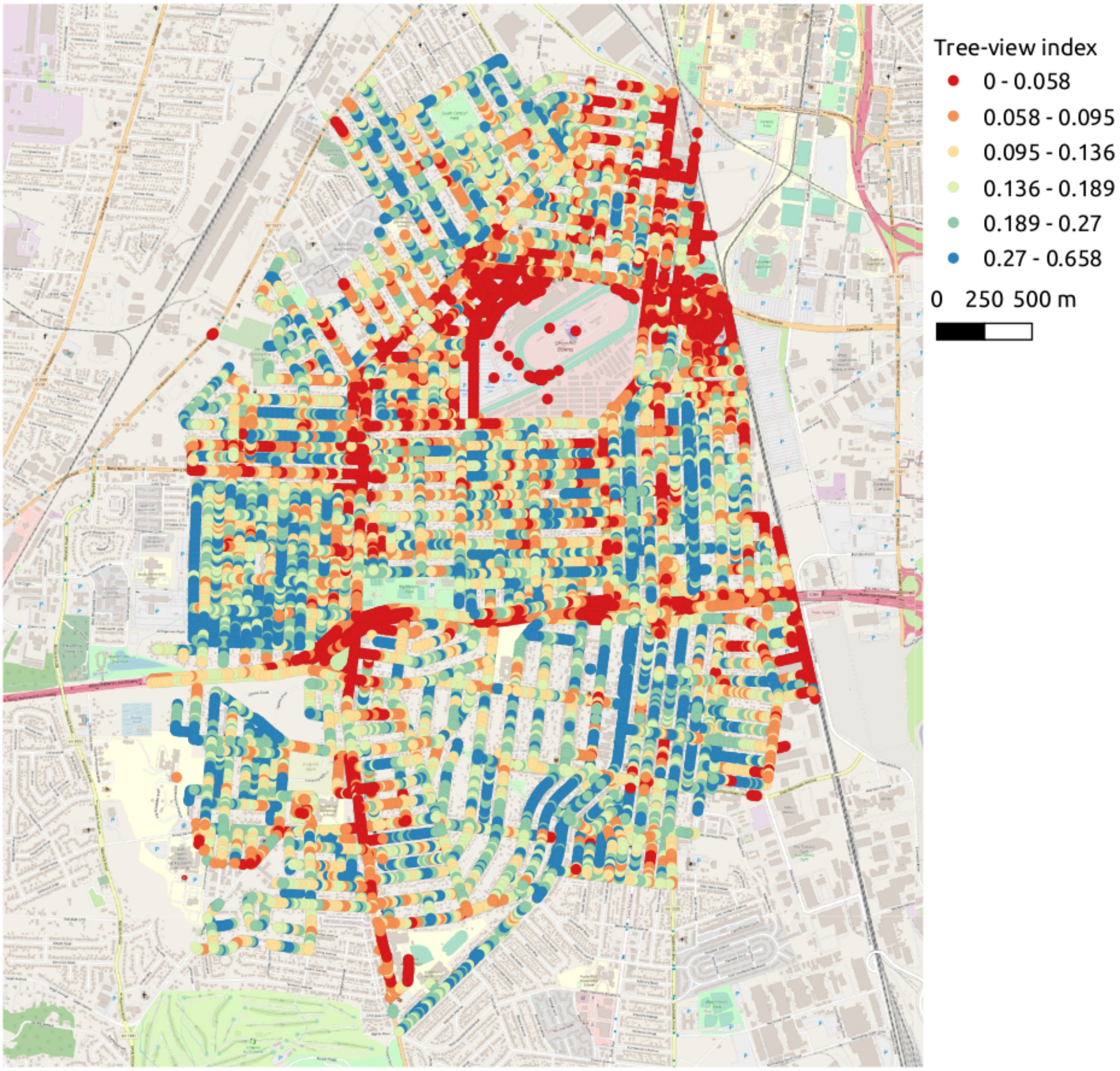


*Figure SI-11: The tree-view index derived from semantic segmentation of Google street view images.*

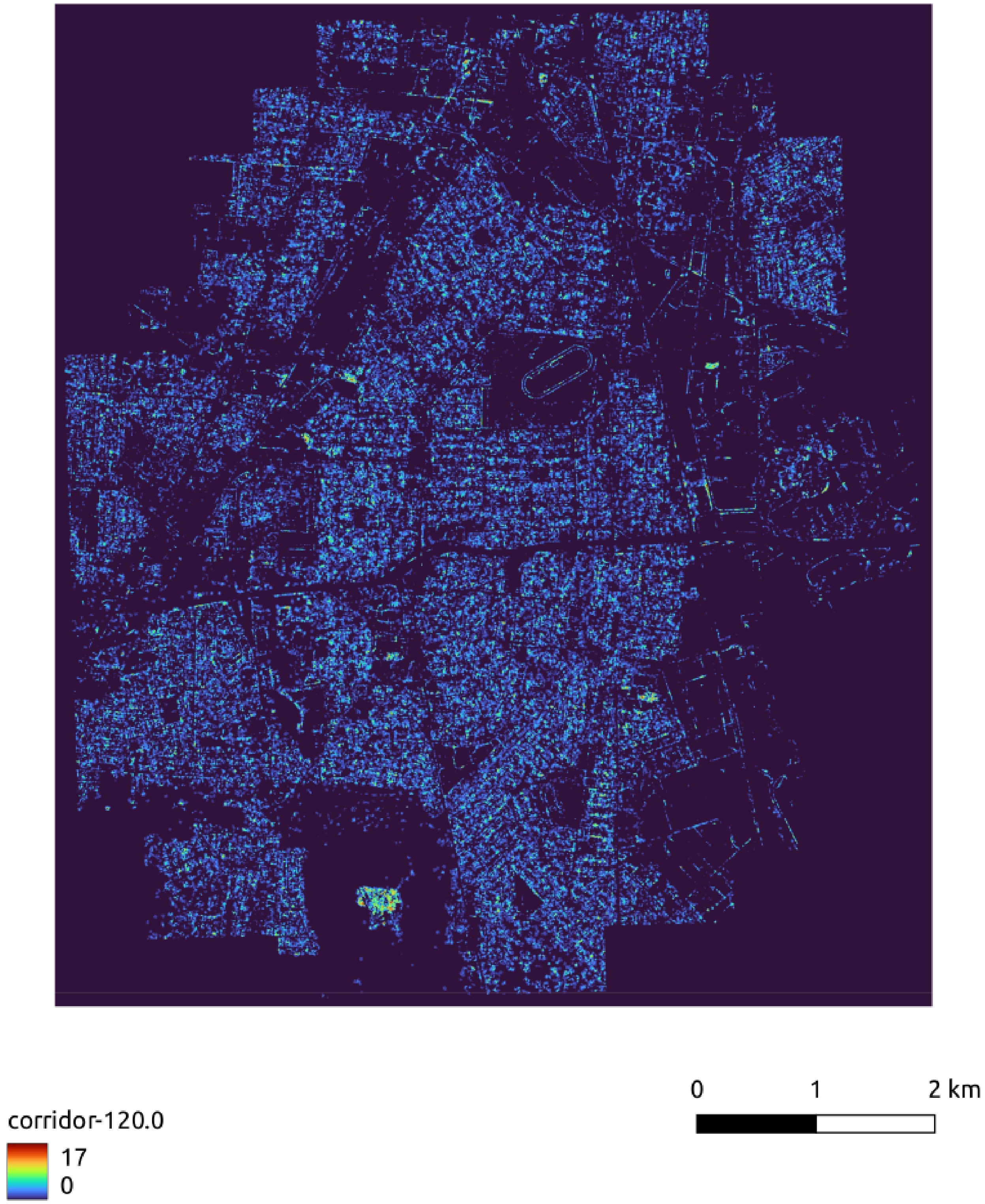


*Figure SI-12: The corridor map generated by Graphab from the canopy patches.*

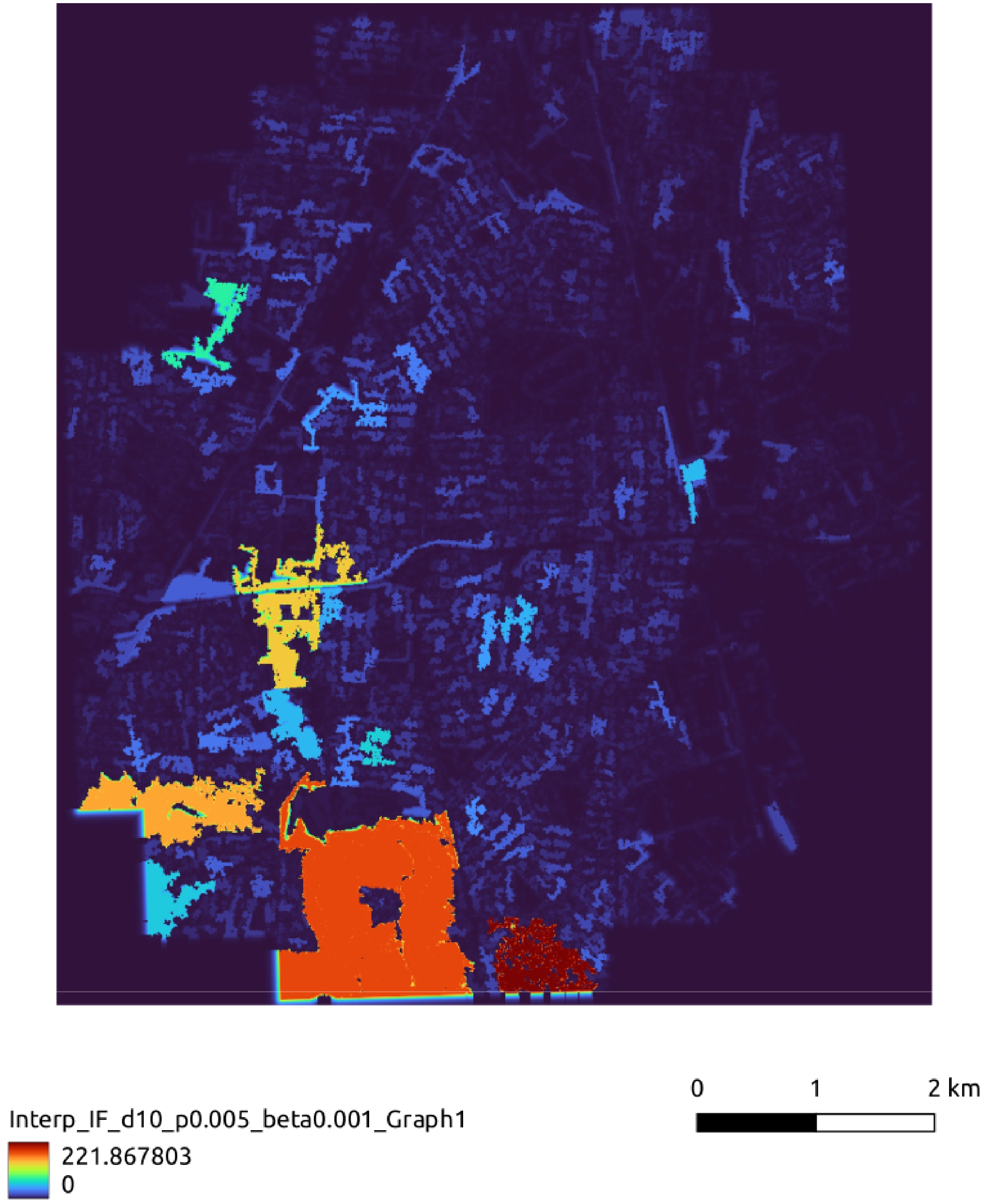

*Figure SI-13: Interaction flux map*

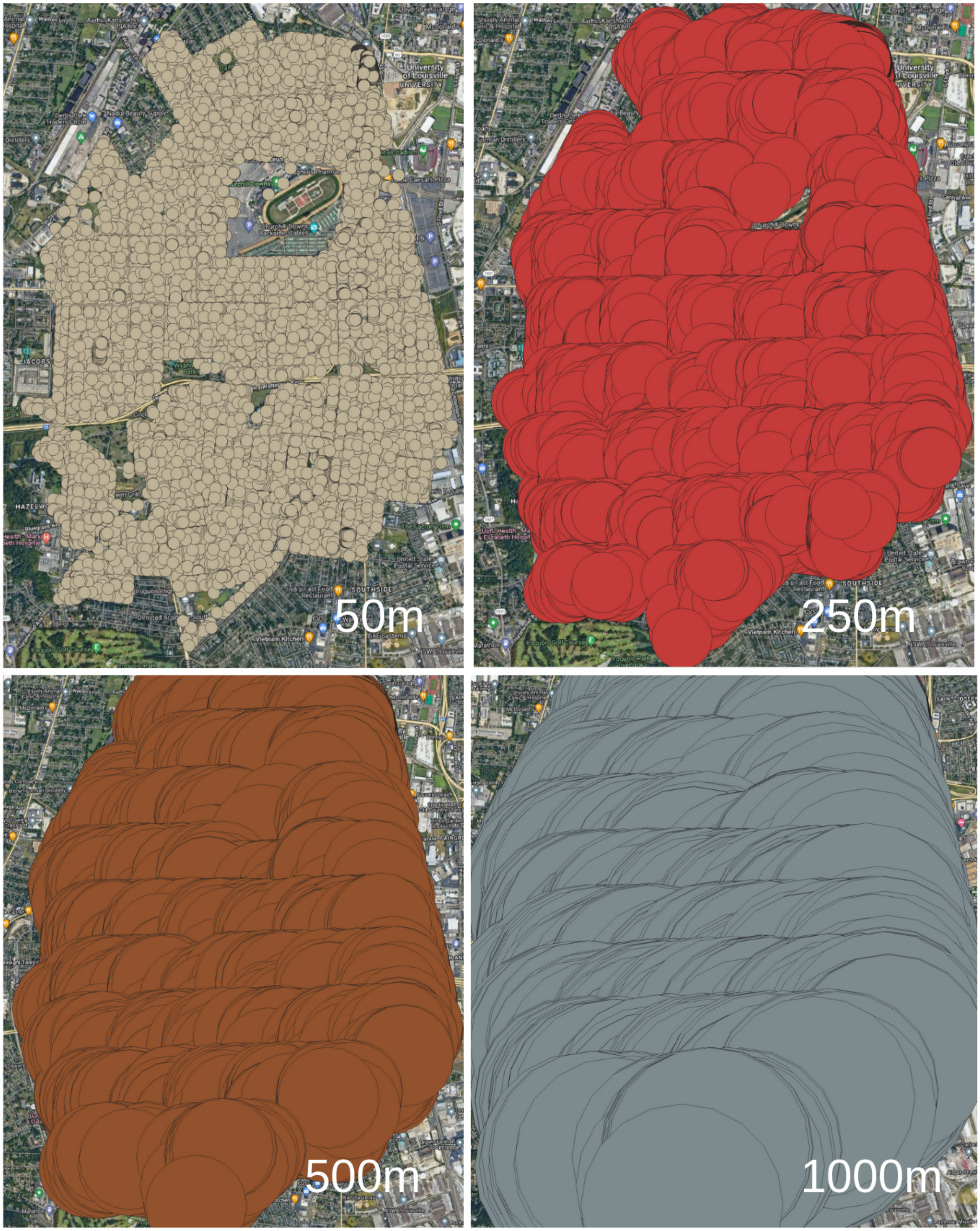


*Figure SI-14: Buffers around each address*

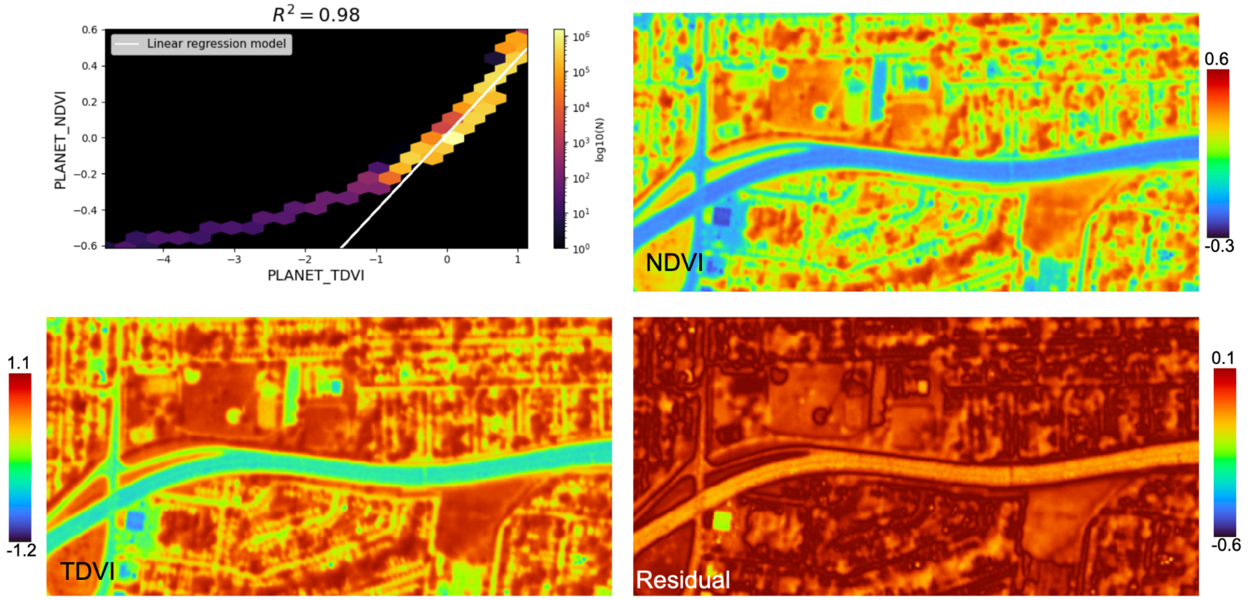


*Figure SI-15: Regressions residuals for TDVI vs NDVI*

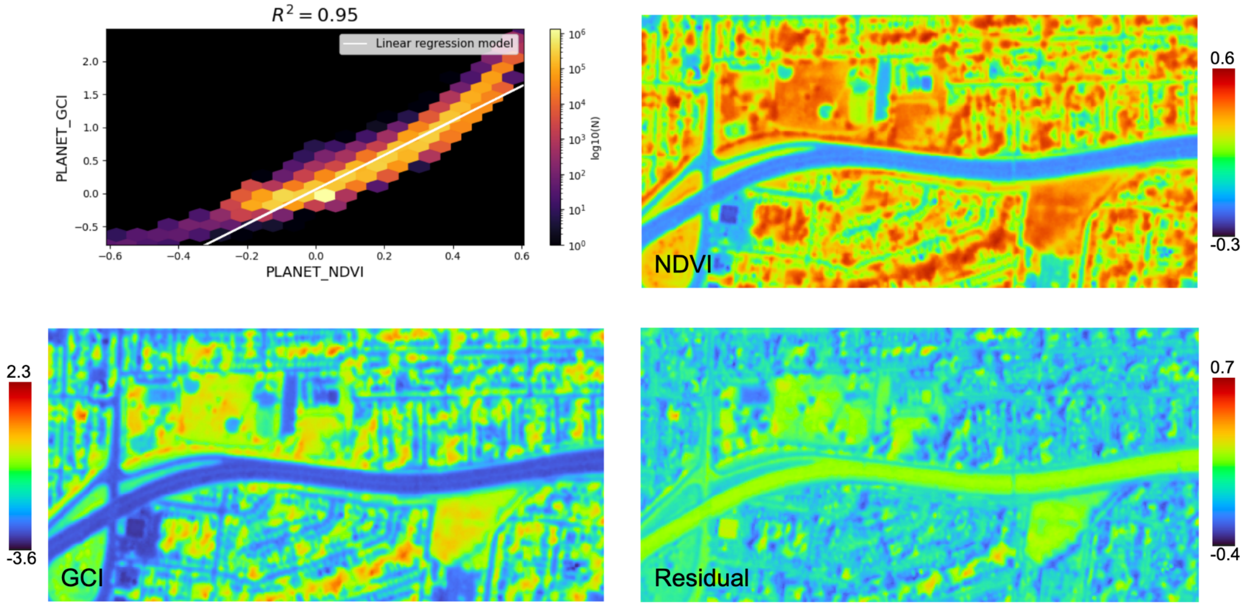


*Figure SI-16: Regressions residuals for GCI vs NDVI- low residual values may be tree shadows.*

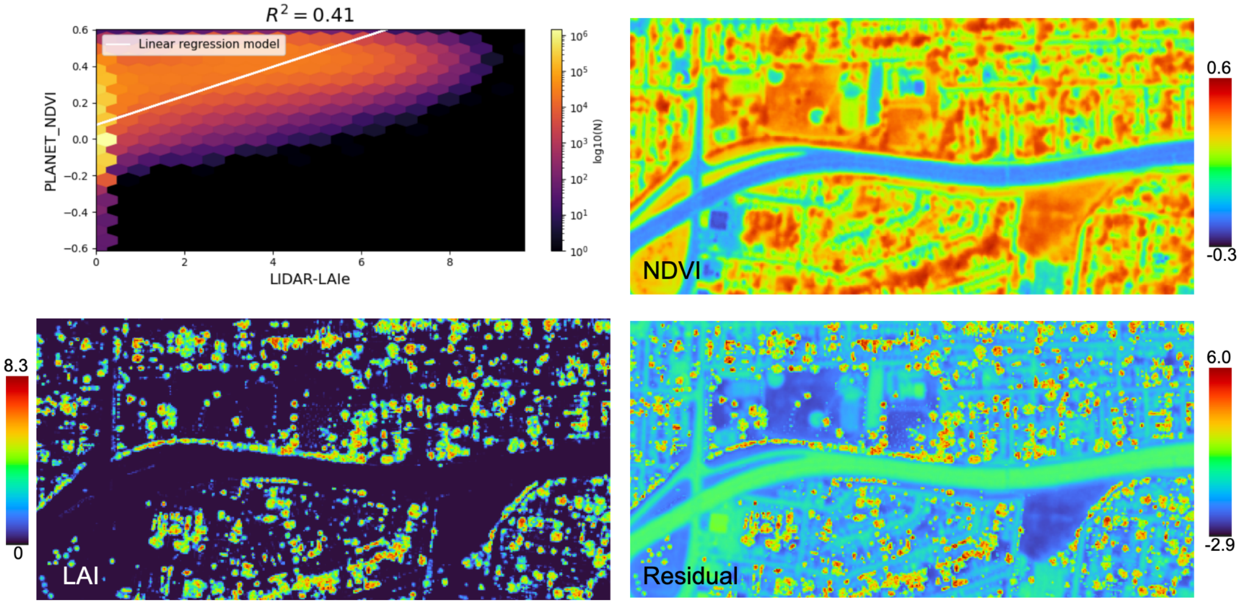


*Figure SI-17: Regressions residuals for LAI vs NDVI*

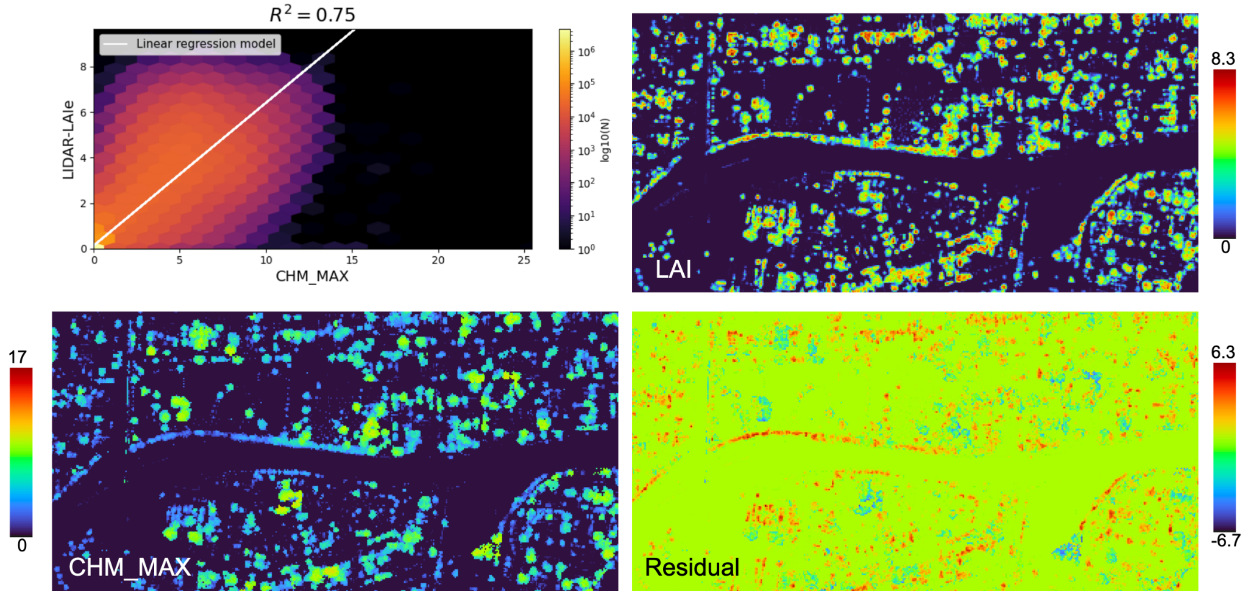


*Figure SI-18: Regressions residuals for Canopy height maximum vs LAI*

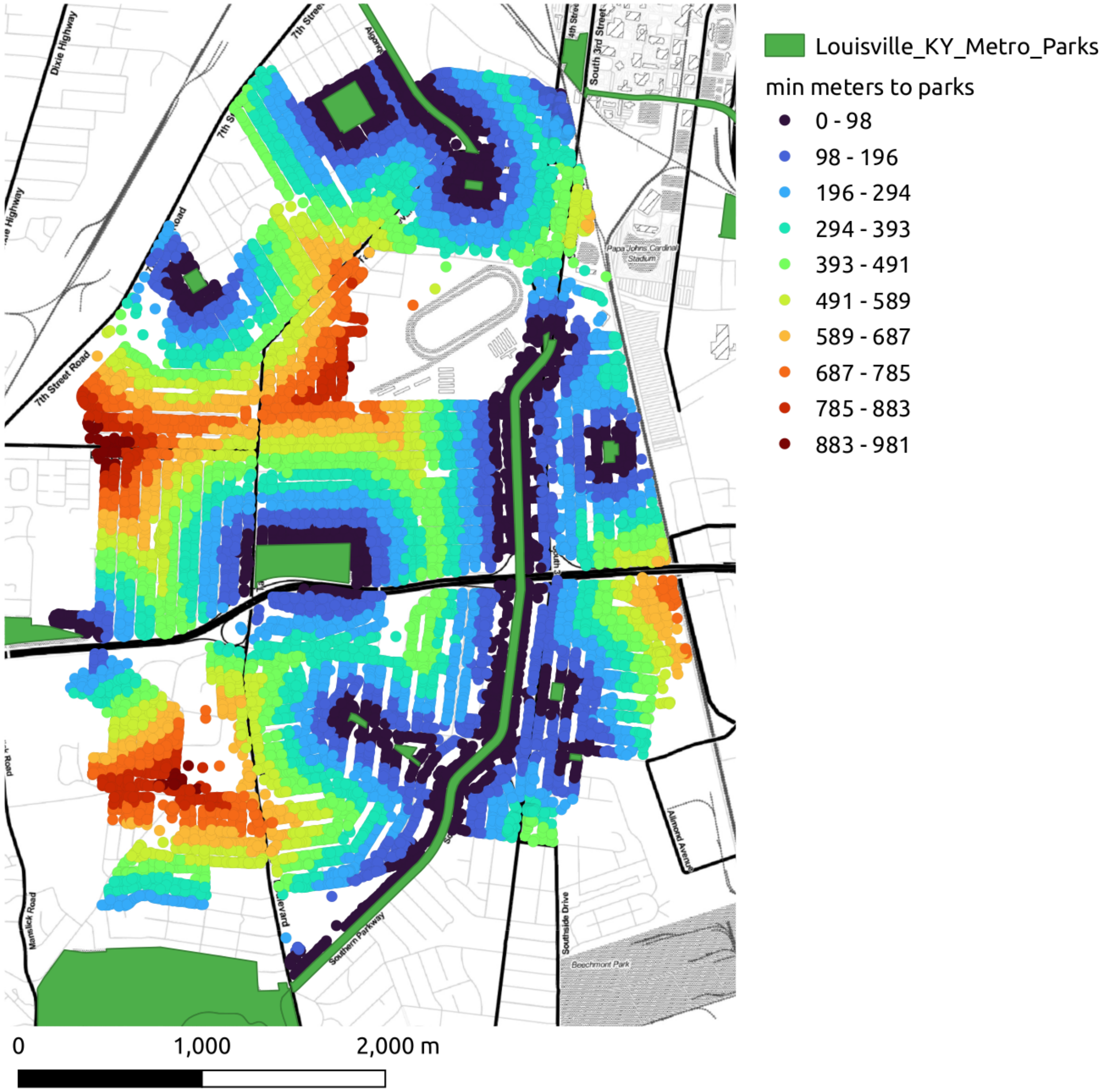


*Figure SI-19: Minimum Distance to parks.*

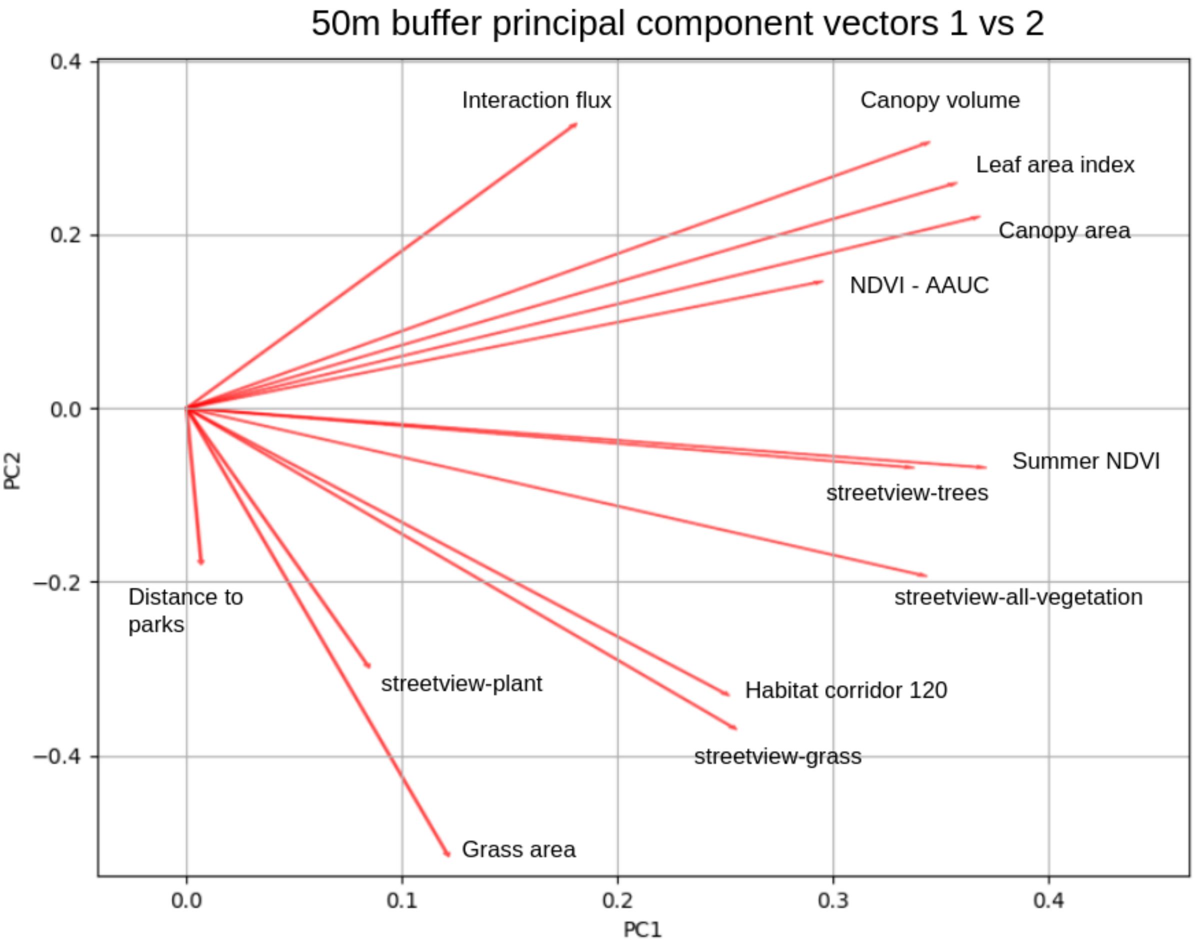


**Figure SI-20:** Principal component vectors 1 and 2 for 50m buffer radii

Figure SI-20 shows a mapping of the component vector ratios between principal components 1 and 2. By vectors in the top of the figure, above the 0 point of the Y axis have a positive relationship between components 1 and two, and includes the interaction flux as the most divergent feature, and a prominent cluster containing canopy volume, area and LAI, along with the annually integrated NDVI area under the curve. The bottom portion of the graph below the zero value of the Y axis contains summer NDVI, streetview trees and streetview-all vegetation appearing near each other, habitat corridors and streetview grass appearing near each other, grass area being fairly isolated, and distance to parks as a low magnitude feature.

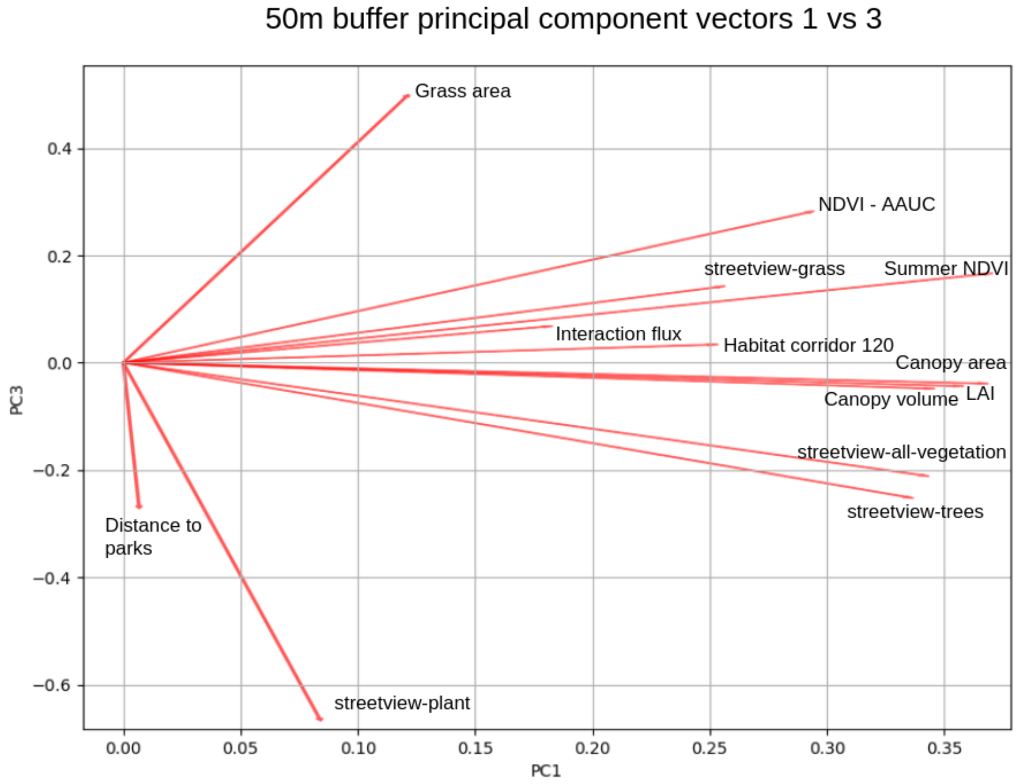


**Figure SI-21:** Principal component vectors 1 and 3 for 50m buffer radii

Figure SI-21 shows a mapping of the component vector ratios between principal components 1 and 3. This suggests grass area, distance to parks and streetview-plant as relatively isolated features, perhaps a cluster containing NDVIs, streetview grass, interaction flux and habitat corriro, another with canopy area, volume and LAI, and another with streetview trees and streetview all vegetation. .

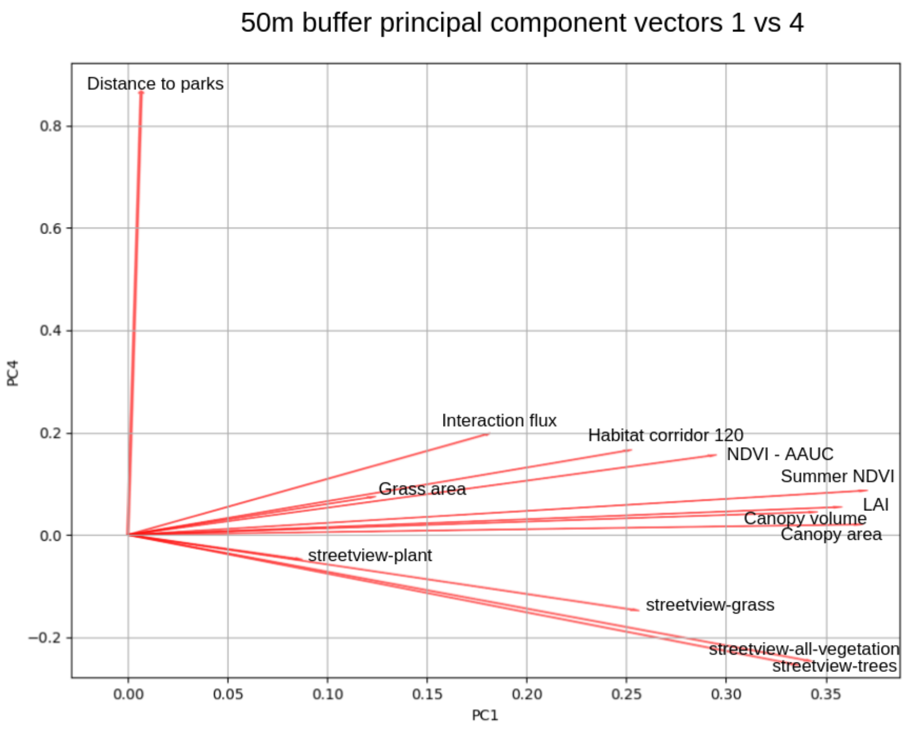


**Figure SI-22:** Principal component vectors 1 and 4 for 50m buffer radii

Figure SI-22 shows a mapping of the component vector ratios between principal components 1 and 4. This suggests distance to parks an isolated feature, perhaps a cluster of streetview metrics, and the rest being a cluster.
